# Direct Oral Anticoagulants vs. Heparin for Cancer-Related Stroke: Augmented Meta-Analysis

**DOI:** 10.1101/2024.11.14.24317340

**Authors:** Muhammed Amir Essibayi, Ahmed Y. Azzam, Ugur Sener, David Altschul, Merve Atik, Zafer Keser

## Abstract

**Background:** Ischemic stroke is common among patients with systemic malignancy, associated with increased risk of neurological deterioration and mortality compared to the general population. Optimal approach to secondary stroke prevention in cancer patients is unclear. In this meta-analysis, we evaluated available data on the use of direct oral anticoagulants (DOACs) and heparin products for stroke prevention in this population.

**Methods:** Using the Nested Knowledge AutoLit software, we performed PubMed search in September 2023 for articles reporting the use of antithrombotics for cancer-associated stroke. We conducted systematic review and meta-analysis. We also used a novel computational augmentation method to amplify the sample size to predict the effect before and after sample size augmentation and predict the results of further trials.

**Results:** Among 253 potential studies screened, 7 were eligible for inclusion. 439 patients were treated with DOACs and 3968 with heparin products. Among patients treated with heparin, intracerebral hemorrhage (8.8 % vs 1.6, p=.02), overall hemorrhagic complications (17.9% vs 3.5%, p<.001), and mortality [28.1% vs 23.5%, p<.001] were respectively significantly higher than those reported among patients who received DOAC for cancer-associated ischemic stroke. No significant difference was observed in the rates of recurrent deep venous thrombosis, clinically significant hemorrhage, and clinical outcomes between the treatment groups. Similar results were shown with augmented meta-analysis.

**Conclusions:** This meta-analysis shows DOACs may have efficacy and safety profile similar to heparin products for recurrent stroke prevention in patients with cancer. Given the small number of studies and limited data, findings should be interpreted with caution.

## INTRODUCTION

Ischemic stroke is a frequent complication of systemic malignancy.(1, 2) Lung, pancreatic, breast, and prostate cancer have been variably associated with increased ischemic stroke risk.(3, 4) Ischemic stroke among patients with cancer tends to be more severe compared to the general population, associated with an increased risk of early neurologic deterioration and in-hospital death.(5, 6) Rate of fatal stroke has been reported as 21.64 per 100,000 person-years among cancer patients, which is two times the risk in the general population.(7)

Several mechanisms can contribute to the development of ischemic stroke in patients with malignancy, including cancer-related hypercoagulability, deep venous thrombosis, pulmonary embolism resulting in stroke in the setting of a right-to-left cardiac shunt, hyperviscosity syndrome in the setting of hematologic malignancies, and direct compression of intracranial vessels from primary or metastatic brain tumors.(2, 8, 9) However, the precise mechanism is frequently unclear, with the event characterized as embolic stroke of undetermined source (ESUS).(3) Emerging data suggest that patients with cancer may represent a distinct subgroup among individuals with ESUS.(3, 10) In a multi-institutional study, mRNA expression profiles from cancer patients who had ischemic stroke demonstrated increased activation of inflammatory, transcription regulation, and hypoxia response pathways compared to stroke patients who did not have a cancer diagnosis.(10) Hypercoagulable state related to cancer may lead to stroke through nonbacterial thrombotic endocarditis (NTE).(11) This is supported by detection of cerebral microemboli in cancer patients with ESUS undergoing transcranial Doppler.(12)

Optimal management of patients with ESUS in the setting of systemic malignancy is unclear.(2) Two large trials, RESPECT-ESUS and NAVIGATE-ESUS, showed no benefit from use of dabigatran or rivaroxaban compared to aspirin for preventing recurrent ischemic stroke in patients with ESUS.(13, 14) However, neither of these trials specifically studied patients with cancer. In a retrospective study of cancer patients with ischemic stroke, there was no difference in recurrent stroke between patients treated with antiplatelet therapy or anticoagulation.(15) Nonetheless, in real-life practice, there is a tendency to utilize anticoagulation when the mechanism of ischemic stroke is attributed to cancer-related hypercoagulability, and enoxaparin was historically the choice of agent due to better safety profile than warfarin in patients with cancer.(16) This led to a design to a pilot randomized trial in which enoxaparin was compared to aspirin in patients who had an ischemic stroke in the setting of malignancy, but enrollment was limited due to injection aversion.(17) In the absence of dedicated studies, direct oral anticoagulants (DOAC) may be considered a reasonable therapy option in cases of ischemic stroke attributed to suspected hypercoagulability. However, safety and efficacy data in patients with cancer and ischemic stroke is limited. In this meta-analysis, we comprehensively reviewed the efficacy of DOAC compared to heparin-based products for secondary stroke prevention in patients with systemic malignancy related ischemic stroke. In addition to conventional meta-analysis, we also used a novel machine learning based statistical technique to augment the sample size included in the meta-analysis. The technique combines standard meta-analysis and augmented meta-analysis.

## METHODS

### Search strategy

Using the Nested Knowledge AutoLit software, we performed a Pubmed literature search in September 2023 for articles reporting on use of antithrombotic therapies to treat of cancer-related ischemic stroke and venous thromboembolism. The search was performed in accordance with PRISMA guidelines. Search strategies were created using a combination of keywords and standardized index terms, including “( ( Cancer AND Ischemic Stroke ) ) AND ( ( Antithrombotic OR Anticoagulation OR Antiplatelet OR Enoxaparin OR Apixaban OR Rivaroxaban OR Dabigatran OR Edoxaban) ) AND ( ( Stroke OR TIA OR Bleeding OR Intracerebral Hemorrhage OR Hemorrhage ) )”. Results were limited to the English language articles.

### Eligibility criteria

Inclusion criteria included primary research studies reporting patients with cancer-related ischemic stroke managed with any oral anticoagulant medication or heparin-based product. Direct oral anticoagulant (DOAC) considered included apixaban, edoxaban, rivaroxaban, and dabigatran. Heparin-based products included low molecular weight heparin and unfractionated heparin. Exclusion criteria included: 1) Editorial or opinion article, 2) Review or secondary article, 3) Case report of <3 patients, 4) In vitro or animal study, and 5) Secondary study of previously reported data.

### Study selection process

Two authors (AME, MA) screened the titles and abstracts for inclusion using Nested Knowledge’s screening software. Full-text articles of the included abstracts were retrieved and screened by the same authors, and all inclusion decisions were approved by senior authors (US, ZK).

### Data extraction and outcome measures

Baseline characteristics of each study population were collected, including age, gender, type of malignancy diagnosed, whether the malignancy was considered active, and presence of systematic metastasis.

Outcomes of interest were stratified by applied medical treatment into two groups: DOAC and Heparin group. Clinical outcomes included a recurrent ischemic stroke or TIA and DVT, any intracranial hemorrhage, and clinically significant other hemorrhagic complications. Clinically significant hemorrhage was defined as any bleeding event that required transfusion of red blood cells, resulted in hospitalization, required surgical intervention, or resulted in death.

Functional outcomes included mRS at 90 days and mortality. Follow-up periods were extracted from each study.

### Risk of bias assessment & Heterogeneity

The risk of bias was assessed among the included studies using the ROBINS-I tool for non-randomized observational studies provided by Cochrane.(18) When possible, sources of heterogeneity among the results of very high heterogeneity (I^2^>80%) were investigated using sensitivity meta-analysis by methods such as removing individual studies or subgrouping by identified potential co-variates.

### Statistical analysis

Continuous variables were reported as mean and standard deviation. Combined means and SDs of included studies were calculated.(19) The statistical difference between baseline patient characteristics pooled from included studies was assessed by comparing the Z score of each group via the “Z Score Calculator for two Population Proportions” open-source tool provided by Social Science Statistics website.(20) The cumulative incidence, defined as event rate per patient at the end of the study was estimated for each study and a 95% confidence interval was calculated. We used a random-effects model to pool incidence rates across studies because of the heterogeneity that we expected in the populations and interventions across various studies. The I² statistic was used to express the proportion of inconsistency not attributable to chance. Subgroup meta-analysis for all outcomes was conducted using Freeman-*Tukey* double arcsine transformation with restricted maximum likelihood (REML) methods and P values were produced by comparing Z scores of arms of interest. The correction factor was null for all events, including zero events.

### Augmentation Meta-analysis

To enhance the statistical power and precision of our meta-analysis, we employed an innovative augmented meta-analysis technique. This method artificially expands the number of included studies by computing averaged values between random pairs of studies from the original dataset. For instance, if our meta-analysis initially contained seven studies, we generated seven extra synthetic studies by randomly choosing two original studies and averaging their results. This would yield a total of fourteen studies in the augmented meta-analysis (the original seven plus seven synthetically produced averages), which bolstered the stability and dependability of the meta-analytic estimates.

Statistical analysis was performed via OpenMeta[Analyst] open-source statistical software and RStudio for the augmentation meta-analysis.

## RESULTS

### Search results & Baseline characteristics

Two-hundred-eighty potential studies were screened. Two-hundred-sixty-one studies were excluded (**Figure 1**). Of these, 97 studied patients with no cancer, 53 were case reports, 60 were review articles, 9 studied primary stroke prevention, 5 were animal studies, 5 did not provide clinical data, 6 did not include antithrombotic therapy, and 3 did not include patients with a stroke diagnosis. Twenty-three studies were excluded due to patients having comorbid atrial fibrillation (AF) diagnosis. Seven studies were eligible for inclusion (**Table 1**).(21–27)

**Figure 1:**
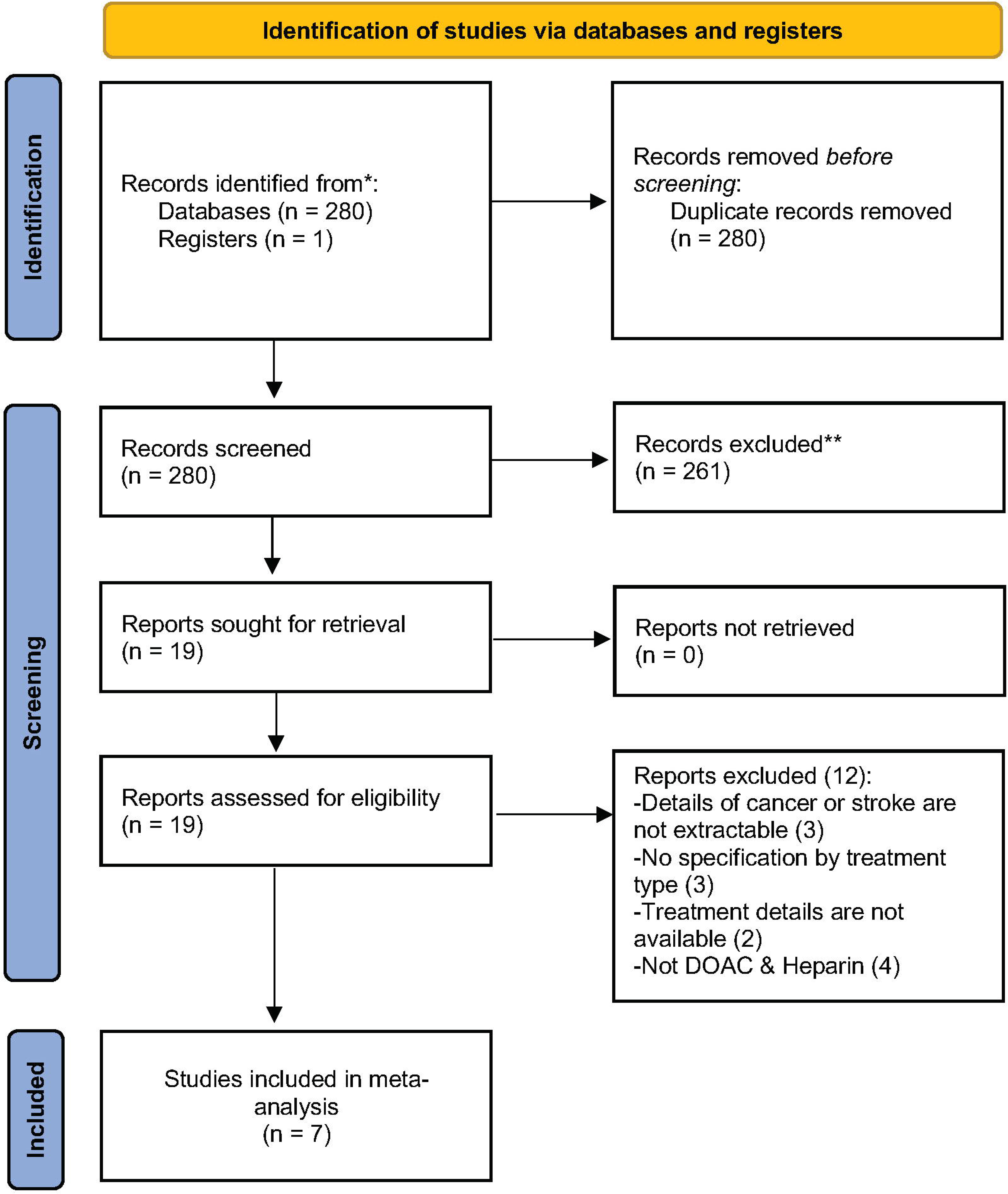
PRISMA flow-diagram

**Table 1:** Details of included studies.

| Author | Year | Ischemia Type | Intervention | Size | Treatment | Timeline of Functional Outcomes | Risk of bias |
| --- | --- | --- | --- | --- | --- | --- | --- |
| Brenner et al. | 2018 | DVT | DOAC | 99 | N/A | 30 days | Low |
|  |  |  | Heparin | 3838 | LWMH |  |  |
| Kawano et al. | 2019 | Stroke | Heparin | 59 | Intravenous unfractionated heparin (19), LWMH (40) | Overall | High |
| Martinez-Majander et al. | 2020 | Stroke | DOAC | 254 | Rivaroxaban (254) | Overall | Low |
| Naito et al. | 2018 | Stroke | DOAC | 2 | Apixaban (1), Edoxaban (1) | Overall | High |
|  |  |  | Heparin | 6 | Subcutaneous unfractionated heparin (5), Intravenous unfractionated Heparin (1) |  |  |
| Nam et al. | 2017 | Stroke | DOAC | 7 | Dabigatran (5), Rivaroxaban (2) | 90 days | Medium |
|  |  |  | Heparin | 41 | LMWH |  |  |
| Weronska et al. | 2021 | DVT | DOAC | 48 | Apixaban (19), Rivaroxaban (16), Dabigatran (13) | Overall | High |
| Yamaura et al. | 2021 | DVT | DOAC | 29 | Edoxaban (24), apixaban (3), Rivaroxaban (2) | 30 days | Medium |
|  |  |  | Heparin | 24 | Unfractionated heparin |  |  |

**Abbreviations:** DOAC, direct oral anticoagulant; DVT, deep venous thrombosis.

While the one study did not specify whether patients have active malignancy or cancer in remission, 5 studies (**Table 2**) clearly defined active cancer as newly-diagnosed cancer, cancer diagnosis within 6 months of stroke, recurrent cancer, metastatic cancer, or cancer requiring ongoing treatment such as chemotherapy, radiotherapy, or surgery. The remaining study relied only on the self-report of the participant of the cancer diagnosis and type.

**Table 2:**
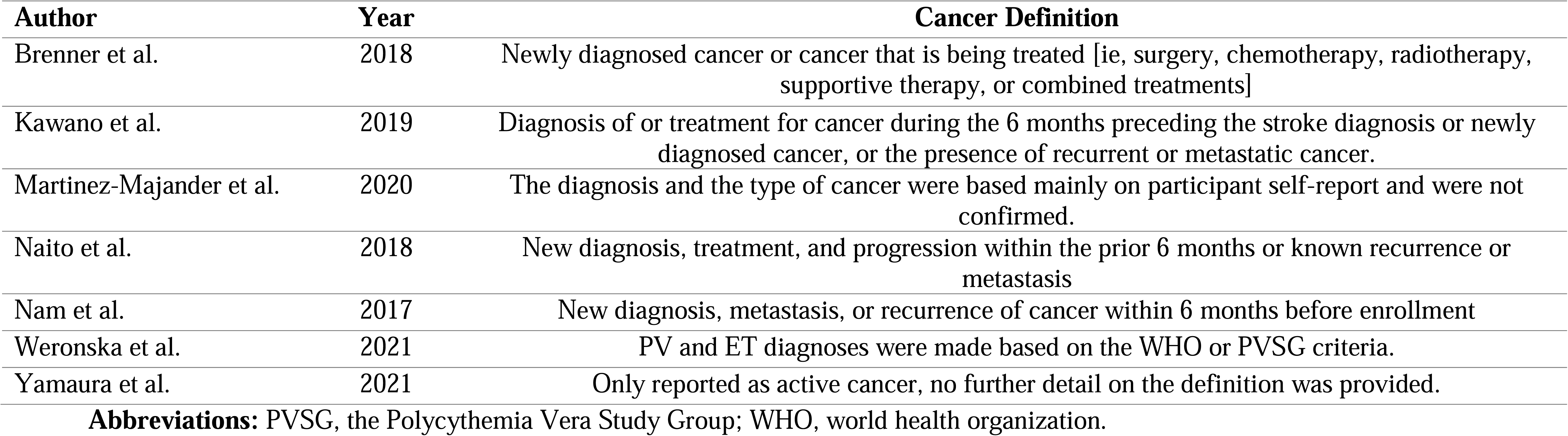
Cancer definition by studies included.

### Direct Oral Anticoagulant therapy

A total of 439 patients with cancer-related ischemic stroke were reported to be treated with a DOAC in 6 studies (**Table 3**). Agents administered included rivaroxaban (n=274, 80.6%), edoxaban (n=25, 7.4%), apixaban in (n=23, 6.7%), and dabigatran (n=18, 5.3%). For 99 patients, specific information about the agent used was not available. Among 86 evaluable patients, mean age was 69. Forty out of 79 (50.6%) evaluable patients were female. Specific cancer diagnosis was available for 86 cases. Among these patients, 49 (57%) had hematologic malignancy. Among solid organ malignancies, lung and hepatobiliary cancers were most common with 12 cases each. Metastasis disease was reported among 30 out of 36 (83.3%) patients. The mean follow-up among 340 patients in 2 studies was 13 ± 8 months.

**Table 3:** Baseline characteristics by treatment.

| **Baseline characteristics (No)** | **Heparin (3968)** | **DOAC (439)** | **Total** | **P value** |
| --- | --- | --- | --- | --- |
| **Age, mean (SD), denominator, year** | 67.1 (12.9), 3968 | 68.9 (6.1), 86 | 67.1 (12.8), 4014 |  |
| **Sex (male), %, denominator** | (50.8), 2511/4939<sup>#</sup> | (49.4), 39/79 | (48.2), 2550/5286 | 0.34 |
| **Cancer type, %, denominator** |  |  |  |  |
| Hematologic | (8.2), 408/4939 | (57), 49/86 | (90.9), 457/5025 | **<.001** |
| Polycythemia Vera | 0 | 34/48 |  |  |
| Essential Thrombocythemia | 0 | 14/48 |  |  |
| Malignant Lymphoma | 0 | 1/29 |  |  |
| Not specified | 408 | 37 |  |  |
| Lung | (13.2), 652/4939 | (13.9), 12/86 | (11.9), 664/5603 | 0.84 |
| Stomach/Esophagus | (13.8), 18/130 | (2.3), 2/86 | (13.3), 20/150 | **0.004** |
| Colorectal | (9.2), 12/130 | (0), 0/86 | (8.5), 12/142 | **0.003** |
| Hepatobiliary | (16.9), 22/130 | (8.3), 12/145 | (20.7), 34/164 | **0.03** |
| Pancreatic | (3.7), 185/4939 | (7), 6/86 | (3.7), 191/5130 | 0.12 |
| Breast | (15.5), 767/4939 | (1.2), 1/86 | (13.5), 768/5707 | **<.001** |
| Genitourinary | (22.3), 1099/4939 | (6), 5/86 | (18.3), 1104/6043 | **<.001** |
| Prostate* | (3.1), 4/130 | (1.2), 1/86 | (7.1) 10/140 | 0.36 |
| CNS tumors | (3.2), 155/4915 | (0), 0/9 | (3.05), 155/5070 | NP |
| **Adenocarcinoma, %, denominator** | (80.8), 72/89 | N/A | (41.7), 72/161 | N/A |
| **Metastatic disease, %, denominator** | (46.3), 2285/4933 | (83.3), 30/36 | (31.9), 2135/7248 | NP |
| **Follow-up, mean (Range), total, month** | 6 (1 – 58.5), 90 | 13 (1 – 30), 340 | 11.2 (10), 430 |  |

\* Prostate cancer was included in the genitourinary cancers but also reported separately for its clinical importance.

The rate of recurrent ischemic stroke or TIA and DVT among patients receiving DOAC was 23.5% (22 out of 263, 95% CI: 0%-53.7%) and 11.3% (14 out of 146, 95% CI: 0%-27.1%), on person-time analysis, 0.4% (95% CI: 0%-0.9%) and 10.7 % (95% CI: 0%-28.5%) per month, respectively. Rate of intracerebral hemorrhage was low among studies; only 4 studies including 86 patients with DOAC reported a rate of intracerebral hemorrhage of 1.6% (2 out of 85, 95% CI: 0%-4.1%). Other clinically significant hemorrhage was reported by six studies among 16 out of 439 patients [3.1% (95% CI: 1.5%-4.7%)]. The overall rate of hemorrhagic complications was 1.4% (14 out of 340, 95% CI: 22.9%-54.8%). Good clinical outcomes, as defined by modified Rankin Score (mRS) of 0-2 were reported among 14 out of 36 patients [38.9% (95% CI: 22.9%-54.8%)]. Twenty-three out of 340 [28.1% (95% CI: 0.6%-55.5%)] died.

### Heparin

A total of 3968 patients with cancer-related ischemic stroke were reported to be treated with heparin products in 5 studies, which included low molecular weighted heparin (LMWH) (n=3919, 98.8%) and unfractionated heparin (n=49, 11.2%). The mean age was 67 and 2428 out of 4939 (49.1%) being female. Genitourinary cancers were also the most commonly reported among 1099 out of 4939 (22.3%) patients, followed by breast (767 out of 4939) and lung (652 out of 4939) cancers. Metastatic disease was reported among 2285 out of 4933 (46.3%) patients. The mean follow-up among 90 patients of 5 studies was 6 ± 14 months. Significant differences between several cancer characteristics of DOAC and heparin were observed (**Table 3**).

The rate of recurrent ischemic stroke or TIA and DVT among patients receiving heparin was 23.5% (30 out of 106, 95% CI: 0%-60%) and 8.4% (362 out of 3862, 95% CI: 4.2%-12.5%), on person-time analysis, 1.1% (95% CI: 0%-2.8%) and 9 % (95% CI: 0%-20.1%) per month, respectively. Reported rate of intracerebral hemorrhage was low among studies; only 3 studies including 71 patients with heparin reported a rate of intracerebral hemorrhage of 8.8% (8 out of 71, 95% CI: 0%-20.4%). Other clinically significant hemorrhage was reported among 195 out of 3909 patients [7.2% (95% CI: 1.3%-13.1%)]. The overall rate of hemorrhagic complications was 17.9% (30 out of 130, 95% CI: 2.4%-33.4%). Good clinical outcomes were reported among 18 out of 65 patients [27.9% (95% CI: 13.1%-42.8%)]. 42 out of 130 [36.5% (95% CI: 12.6%-60.4%)] evaluable patients died during the reported follow up periods. Among patients treated with heparin, intracerebral hemorrhage, overall hemorrhagic complications, and mortality were significantly higher (p<.05) than those reported among patients who received DOAC for cancer-related ischemic stroke (**Table 4**). Forest plots of meta-analysis outcomes were available in **Figure 2**.

**Table 4:** Treatment outcome.

| **Treatment** | **Heparin, % (%95 CI)** | **DOAC, % (%95 CI)** | **P value** |
| --- | --- | --- | --- |
| **Recurrent ischemic stroke or TIA** | 28.2 (0.0-60), 30/106 | 23.5 (0.0-53.7), 22/263 | **<.00001** |
| **Recurrent ischemic stroke or TIA (Person-time analysis)** | 9 (0.0-20.1)/month | 0.4 (0.0-0.9)/month | **<.00001** |
| **Recurrent DVT** | 8.4 (4.2-12.5), 362/3862 | 11.3 (0.0-27.1), 14/176 | 0.63 |
| **Recurrent DVT (Person-time analysis)** | 1.1 (0.0-2.8)/month | 10.7 (0.0-28.5)/month | **<.00001** |
| **Clinically significant hemorrhage** | 7.2 (1.3-13.1), 195/3909 | 3.1 (1.5-4.7), 16/439 | 0.24 |
| **Intracerebral hemorrhage** | 8.8 (0.0-20.4), 8/71 | 1.6 (0.0-4.1), 2/86 | **0.02** |
| **Overall hemorrhagic complications** | 17.9 (2.4-33.4), 30/130 | 3.5 (1.1-6.0), 14/340 | **<.00001** |
| **Good clinical outcome (mRS 0-2)** | 27.9 (13.1-42.8), 18/65 | 38.9 (22.9-54.8), 14/36 | 0.25 |
| **Mortality** | 36.5 (12.6-60.4), 42/130 | 28.1 (0.6-55.5), 23/340 | **<.00001** |

**Abbreviations:** CI, confidence interval; DOAC, direct oral anticoagulant; DVT, deep venous thrombosis; mRS, modified Rankin scale; TIA, transient ischemic stroke

**Figure 2A:**
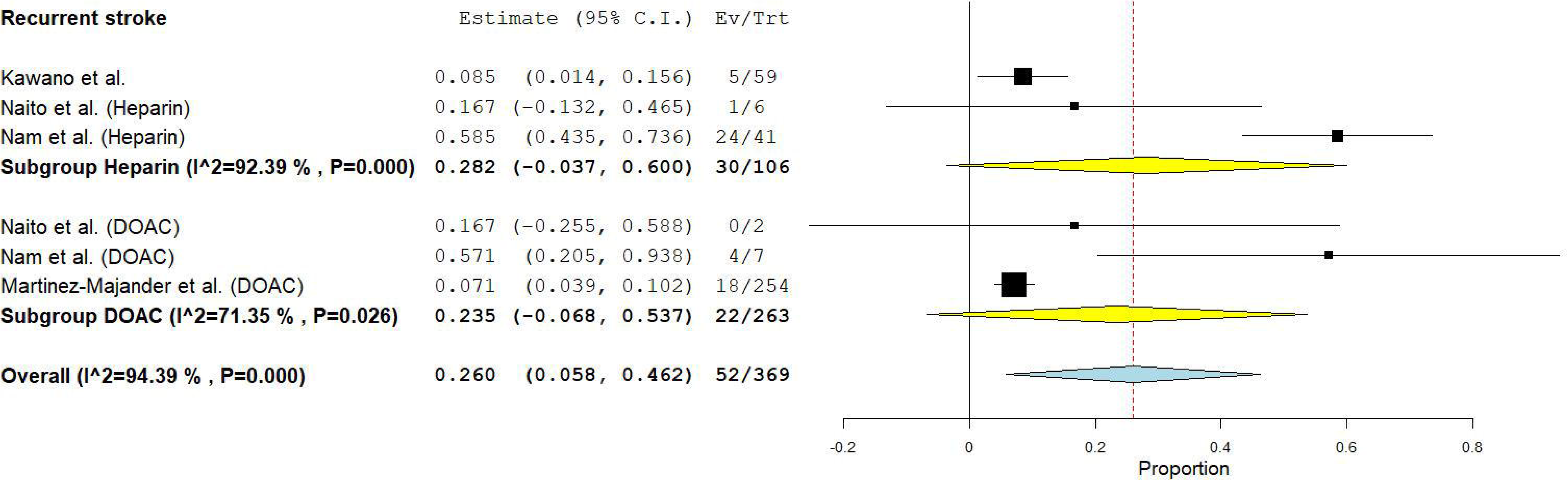
Recurrent stroke

**Figure 2B:**
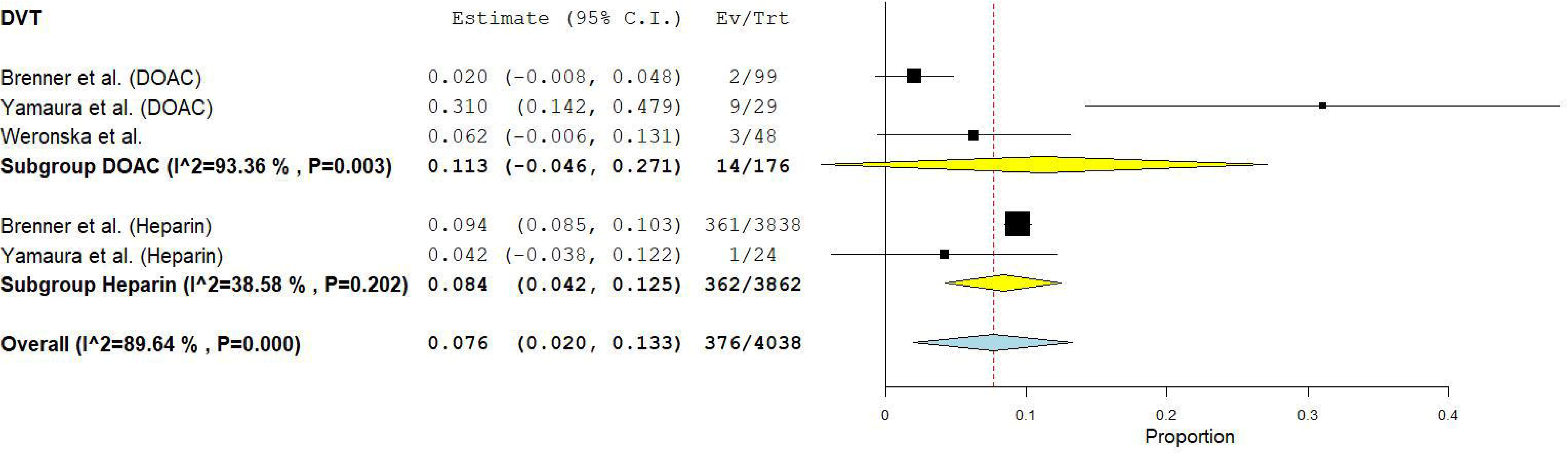
Deep venous thrombosis (DVT)

**Figure 2C:**
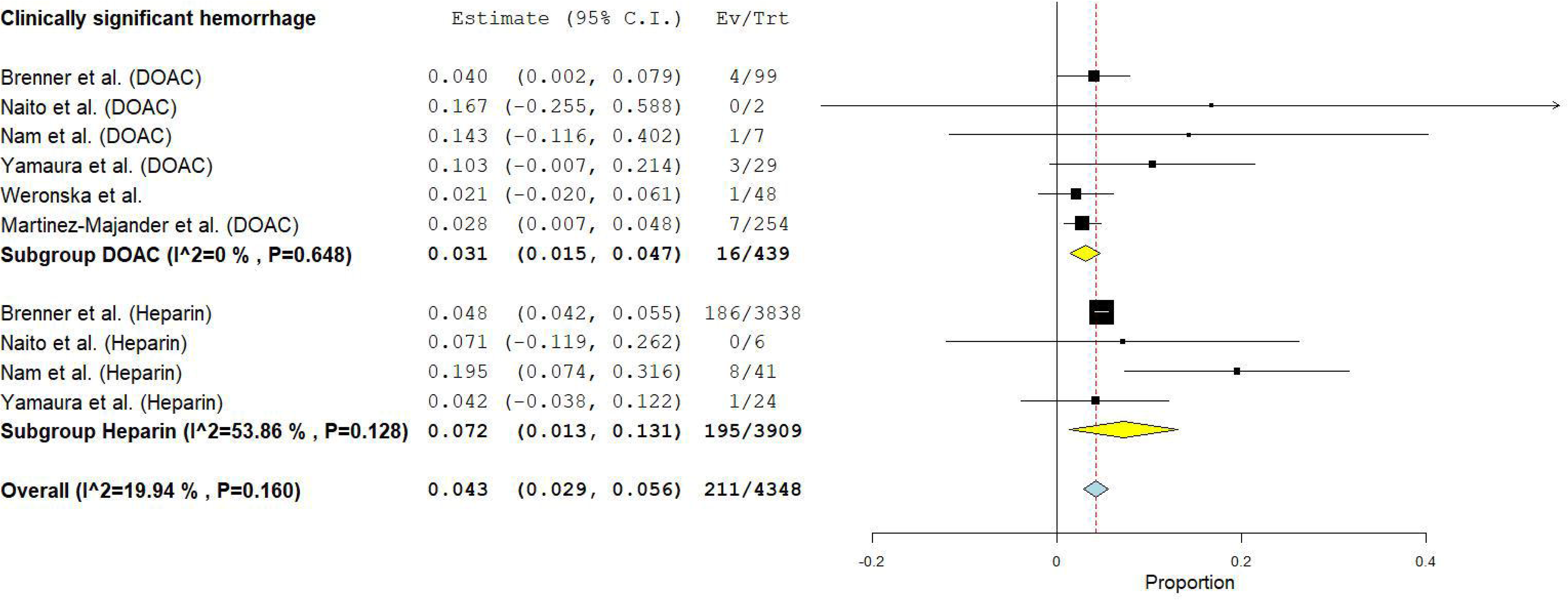
Clinically significant hemorrhage

**Figure 2D:**
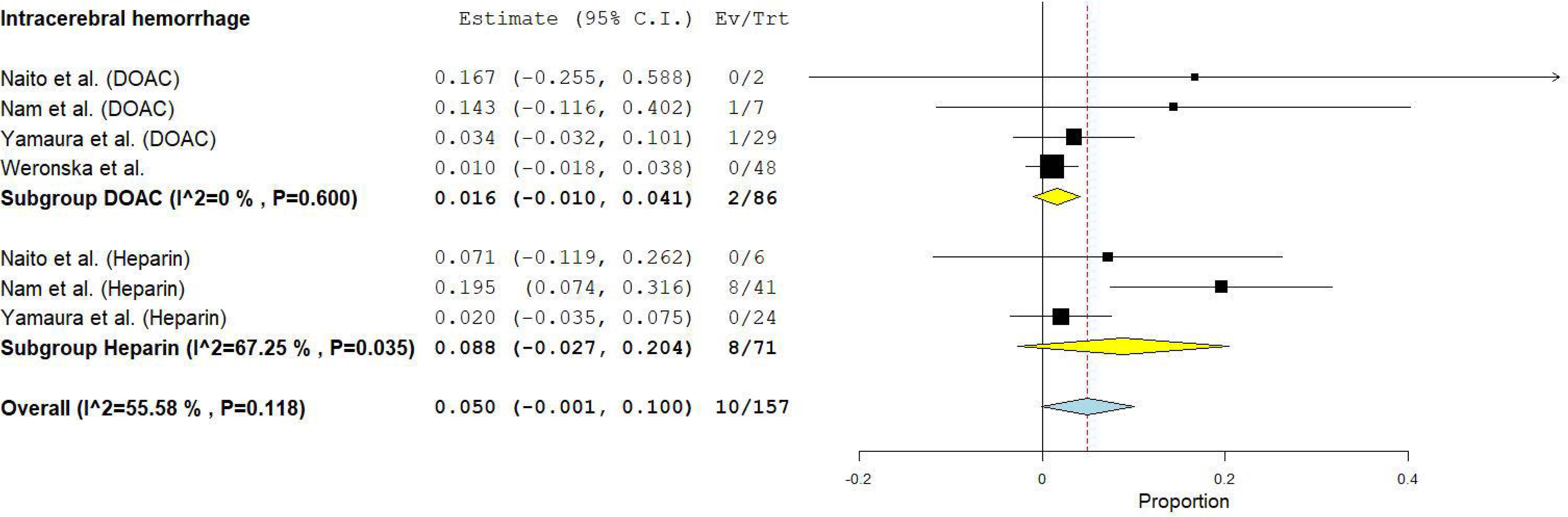
Intracerebral hemorrhage

**Figure 2E:**
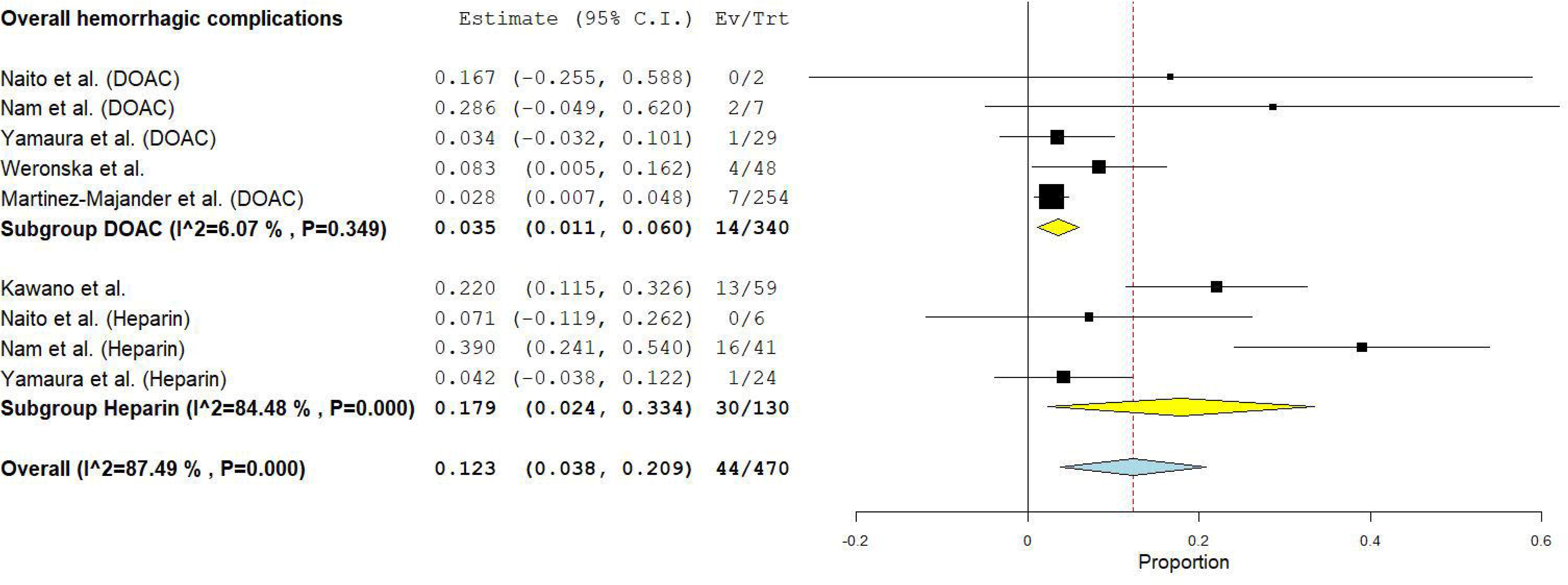
Overall hemorrhagic complications

**Figure 2F:**
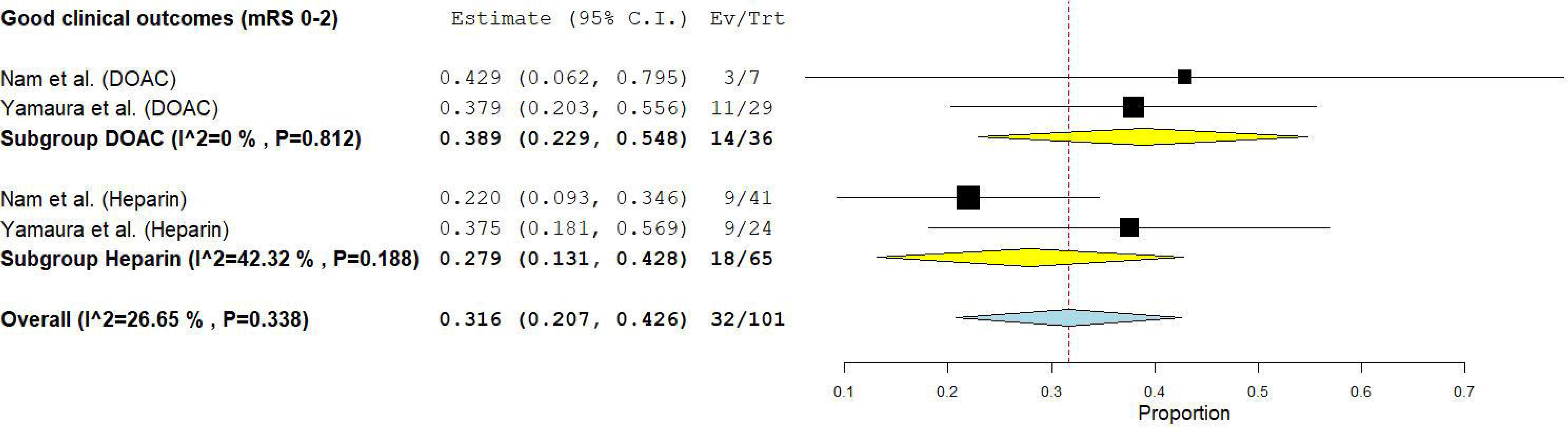
Good clinical outcomes

**Figure 2G:**
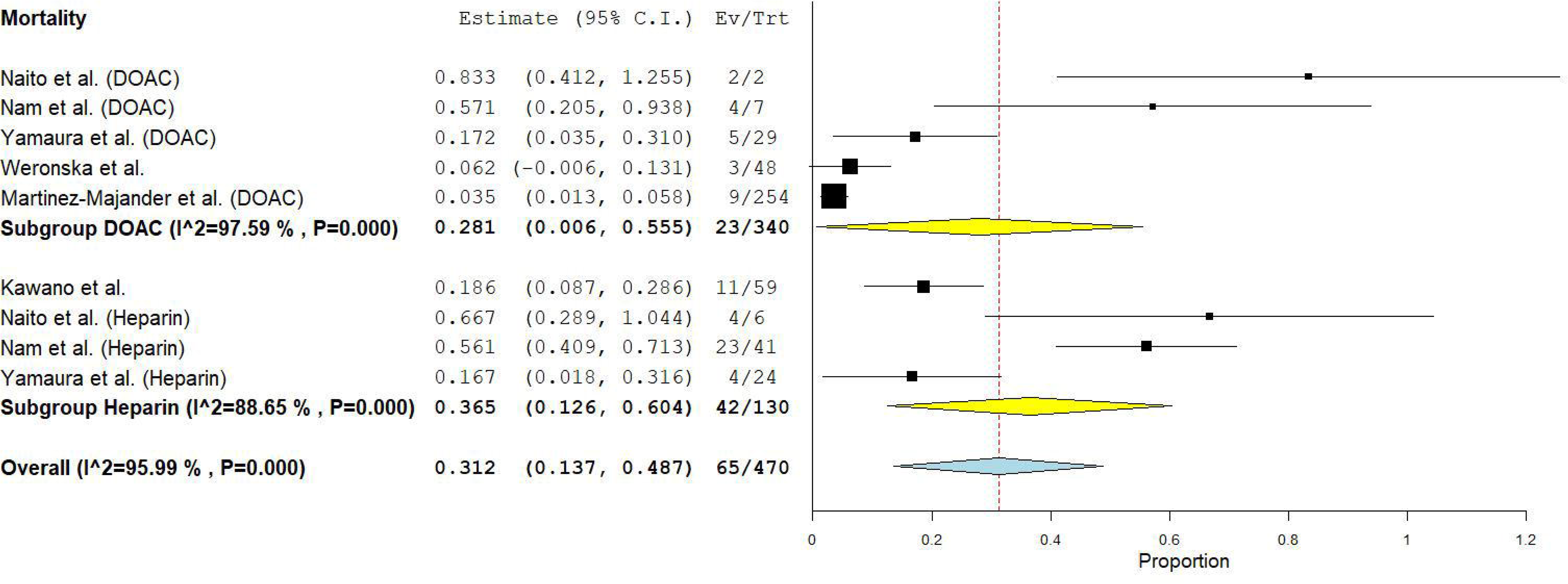
Mortality

### Sensitivity analysis

Very high (I^2^≥ 80) heterogeneity was noted among several outcomes including recurrent/de novo stroke, clinically significant hemorrhage, overall hemorrhagic complications, and mortality. Sensitivity analysis by removing individual studies with low weight in the meta-analysis was performed, however, I^2^ remained high among the corresponding outcomes. Due to the small study number included in the study and by each group, stratification by co-variates of potential impact was inapplicable. The high heterogeneity was attributed to the notable difference in cancer types and heterogeneous follow-ups among treatment groups.

### Augmentation Meta-analysis

An augmented meta-analysis was performed, during which seven synthetic studies were added to all targeted outcomes. The number of synthetic studies generated for each outcome was chosen to match the number of original studies for that outcome. For example, if there were seven original studies for an outcome, seven synthetic studies were added. This allowed the synthetic studies to complement the original studies. For subgroups with only two original studies, just one synthetic study was added by averaging those two studies. Adding only one synthetic study was necessary in those cases to avoid overrepresenting the limited data. The generated results are possible theoretical data points within the dimensions of the original data points of the outcomes. This hypothesis is illustrated in **Figure 3A**. In **Figure 3B**, the black dots represent an example of the original data points extracted in our analysis, plotted on a two-dimensional graph. The blue dots represent synthetically generated data points occupying the space surrounded by the original black points. Connecting the black points shows the blue points falling within the range of the original data. This demonstrates that the synthetic data points are possible outcomes derived from the original results and can be used to augment the analysis. This concept was also demonstrated in **Figure 4A** and **Figure 4B**.

**Figure 3A:**
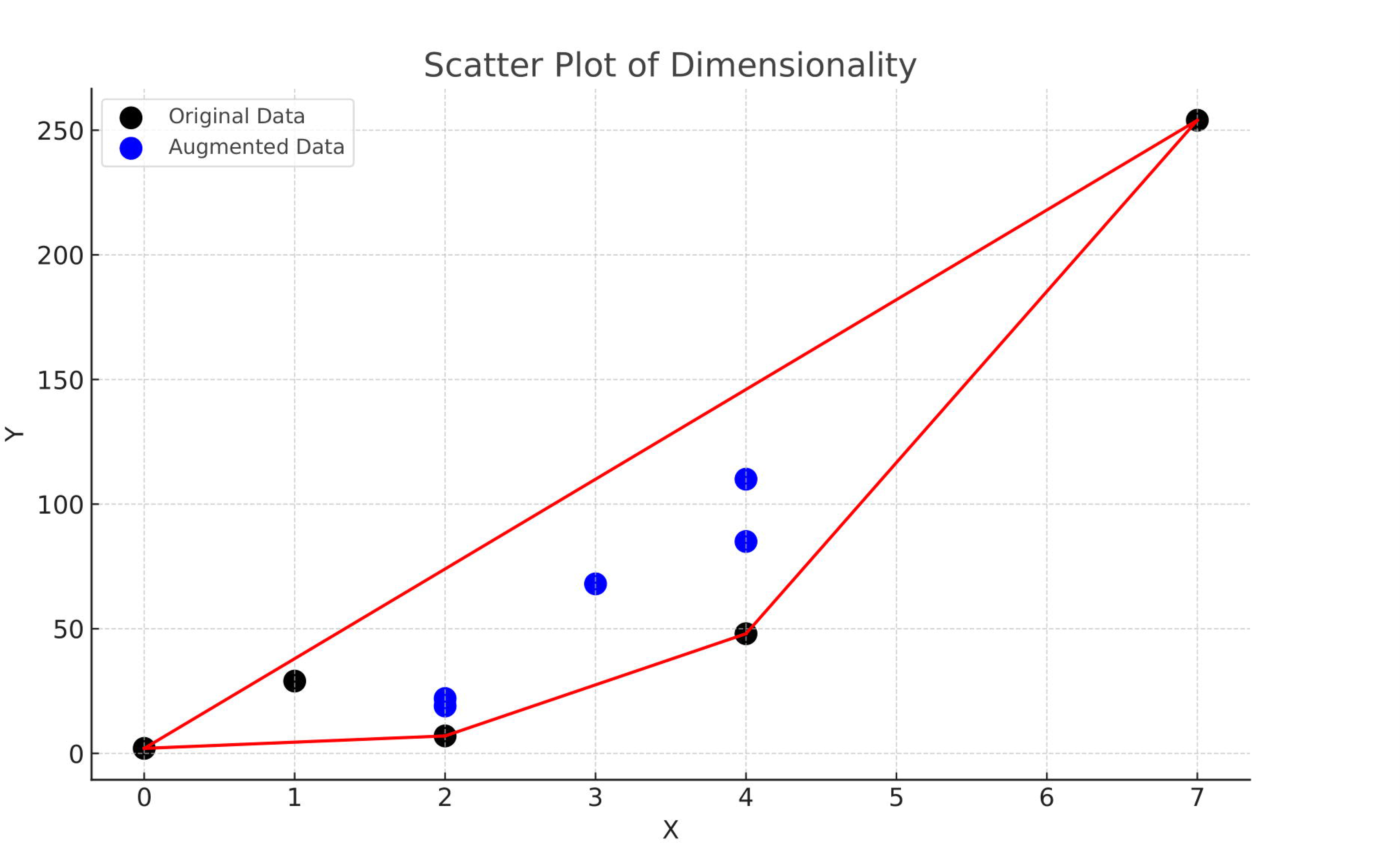
Hypothesis illustration.

**Figure 3B:**
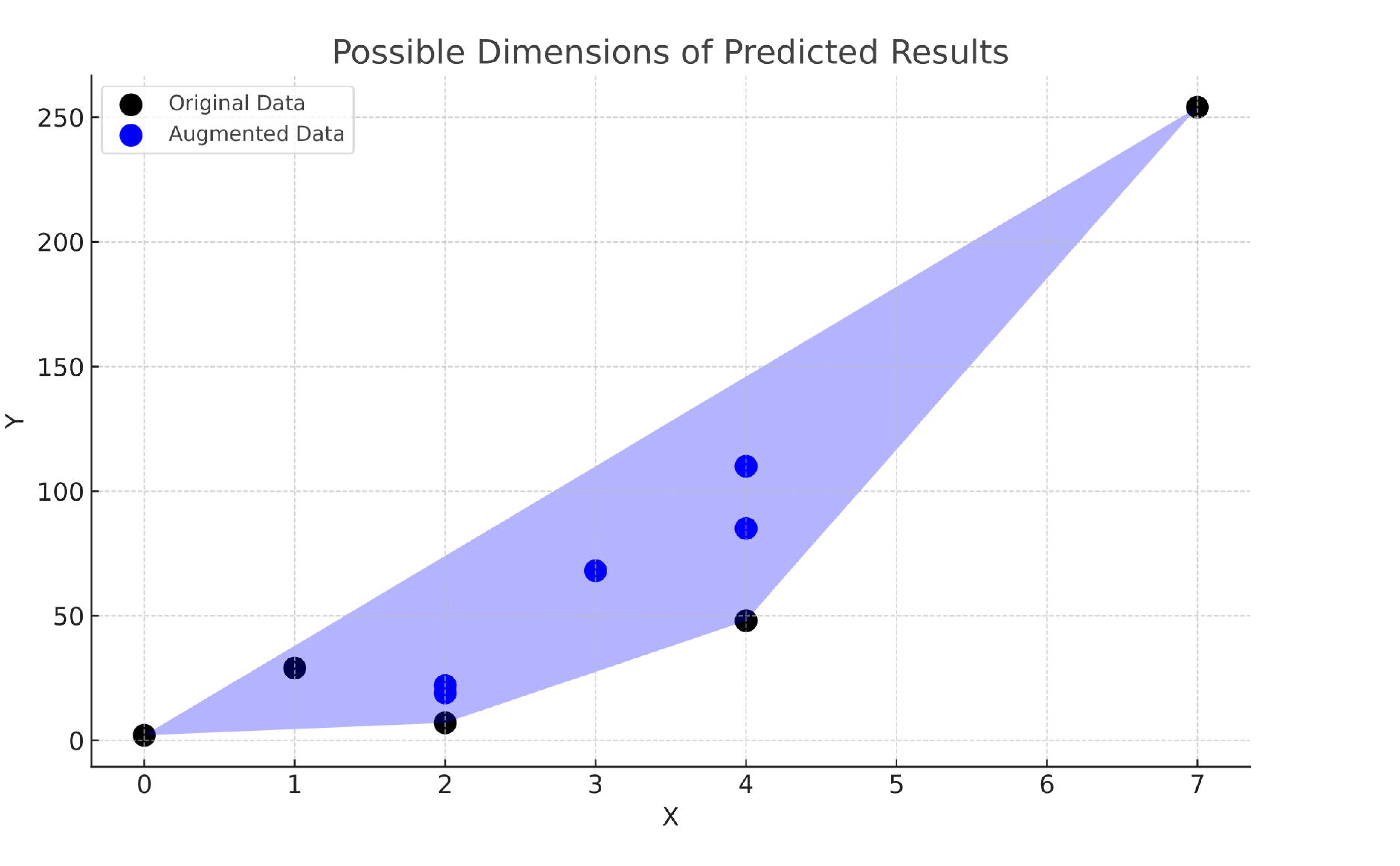
Original data points (black dots) and augmentation-generated plots (blue points) in two-dimensional space.

**Figure 4A and Figure 4B:**
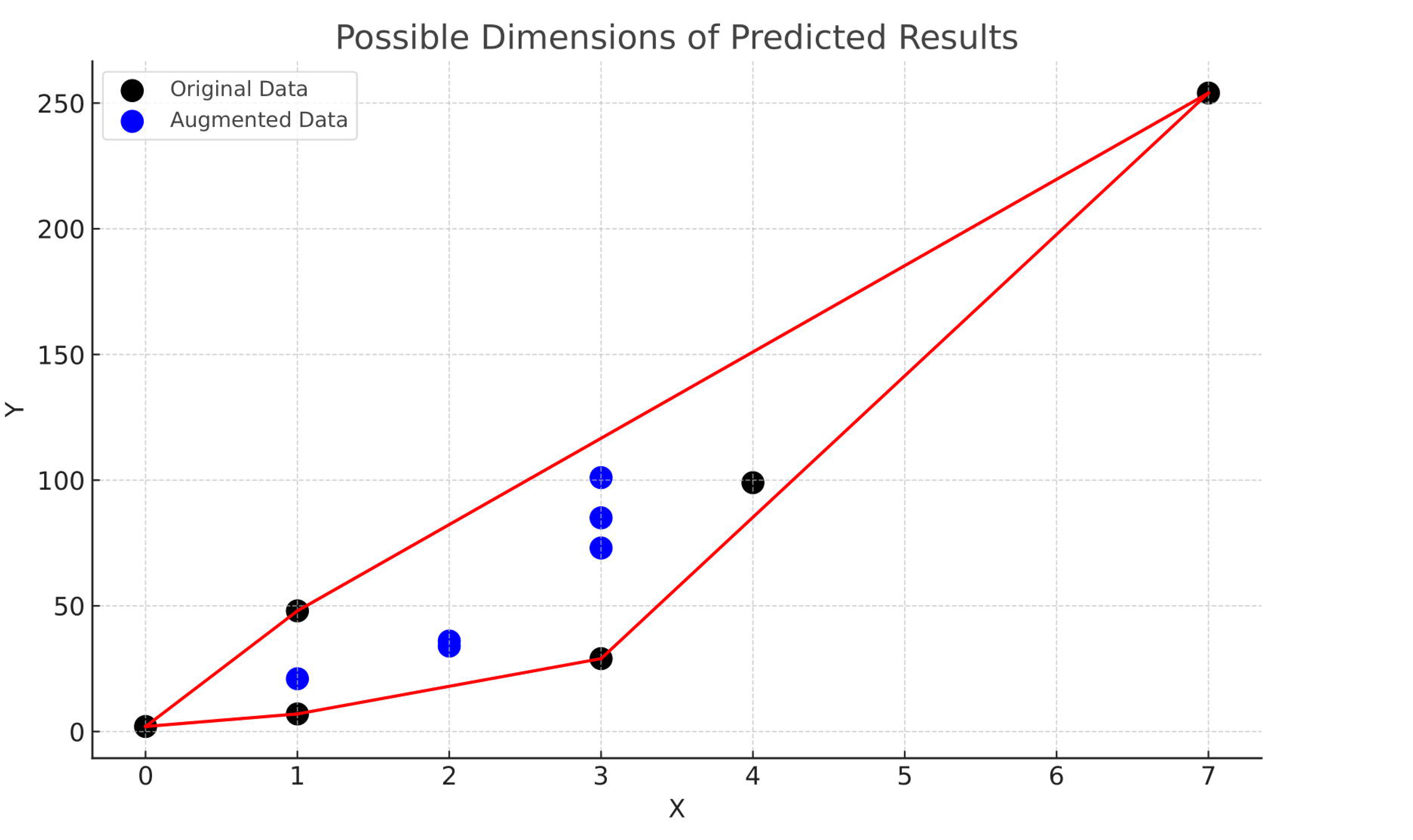
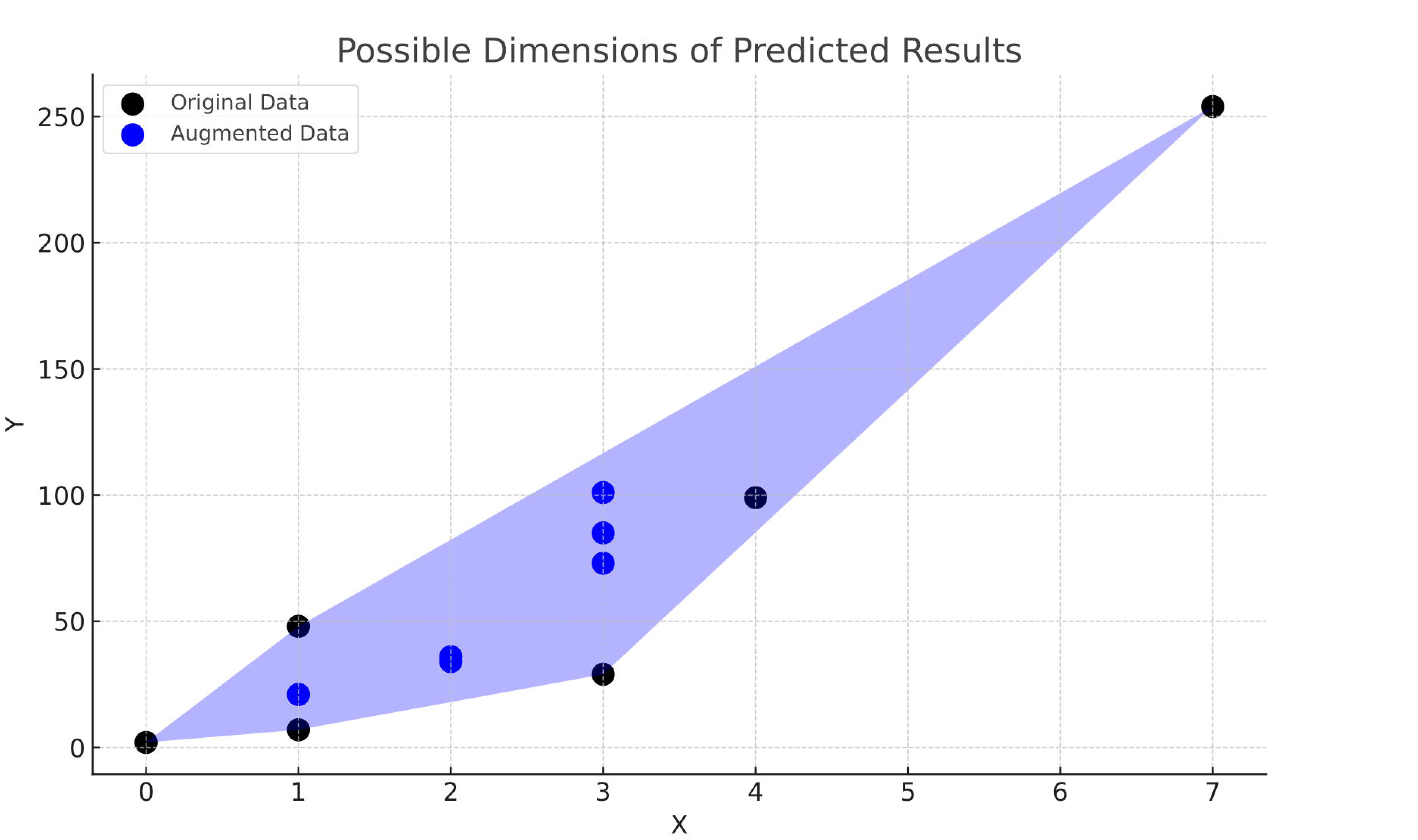
Demonstration of the concept.

The recurrent stroke rate after augmentation was 7.0% (95% CI: 6-10%) with DOAC versus 49.0% (95% CI: 13-86%) with heparin. High heterogeneity existed for both groups (DOAC: I2=68%, p<0.01; heparin: I2=82%, p<0.01). Sensitivity analysis resolved the heterogeneity (DOAC: I2=0%, p=0.69; heparin: I2=67%, p=0.05) (**Figure 5A**). The recurrent stroke rate was lower with DOAC (7.0%, 95% CI: 5-9%) compared to heparin (9.0%, 95% CI: 5-13%), with the difference being marginally significant (p=0.049) (**Figure 5B**).

**Figure 5A:**
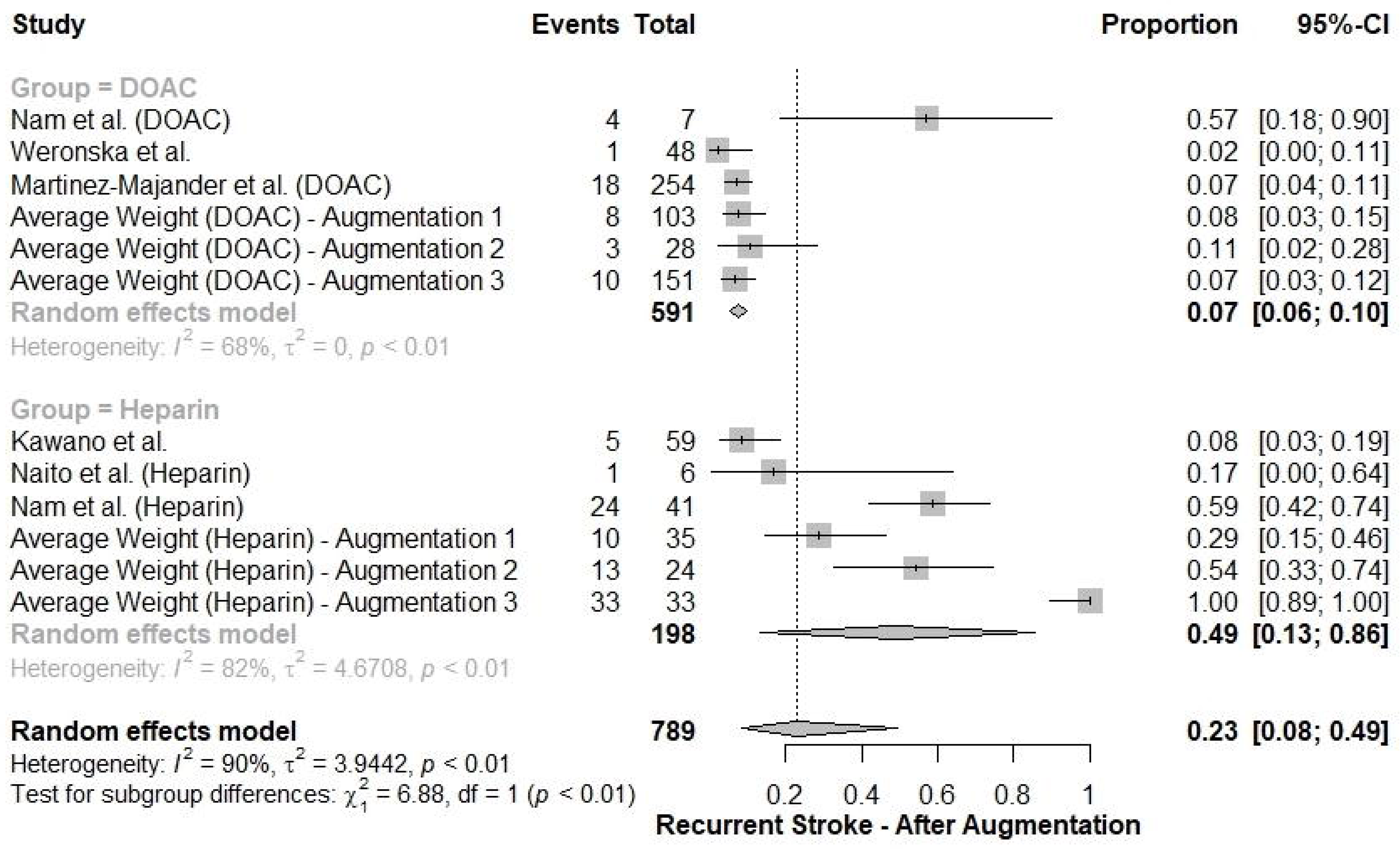
Sensitivity analysis to resolve heterogeneity in recurrent stroke rates between DOAC and heparin groups after augmentation.

**Figure 5B:**
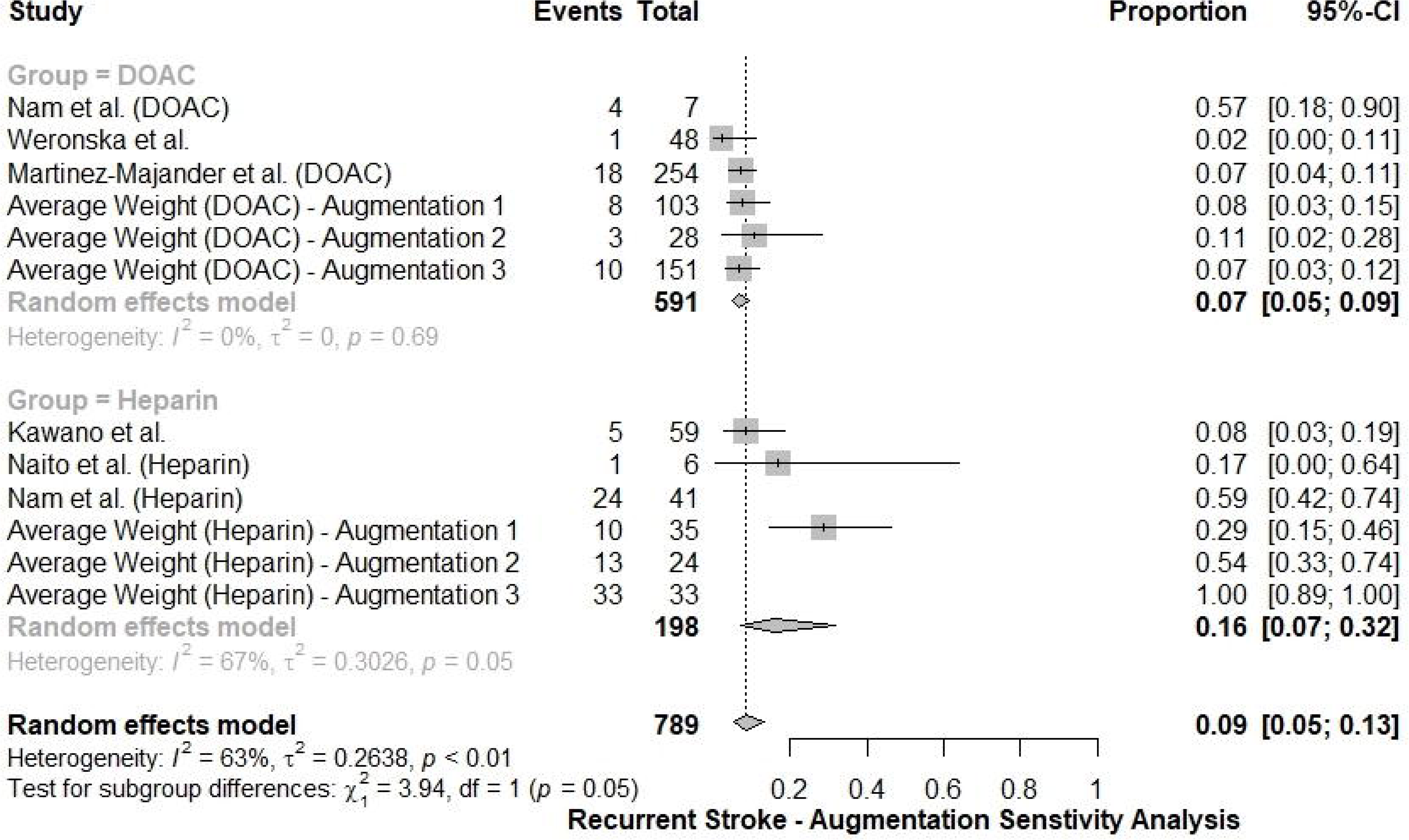
Recurrent stroke rates for DOAC versus heparin after conducting sensitivity analysis.

The DVT rate after augmentation was 10.0% (95% CI: 7-13%) with DOAC versus 9.0% (95% CI: 9-10%) with heparin. High heterogeneity existed for DOAC (I2=62%, p<0.01). Sensitivity analysis resolved the DOAC heterogeneity (I2=28%, p=0.21) (**Figure 6A**). The DVT rates were comparable between DOAC (8.0%, 95% CI: 5-13%) and heparin (9.0%, 95% CI: 9-10%) (p=0.48) (**Figure 6B**).

**Figure 6A:**
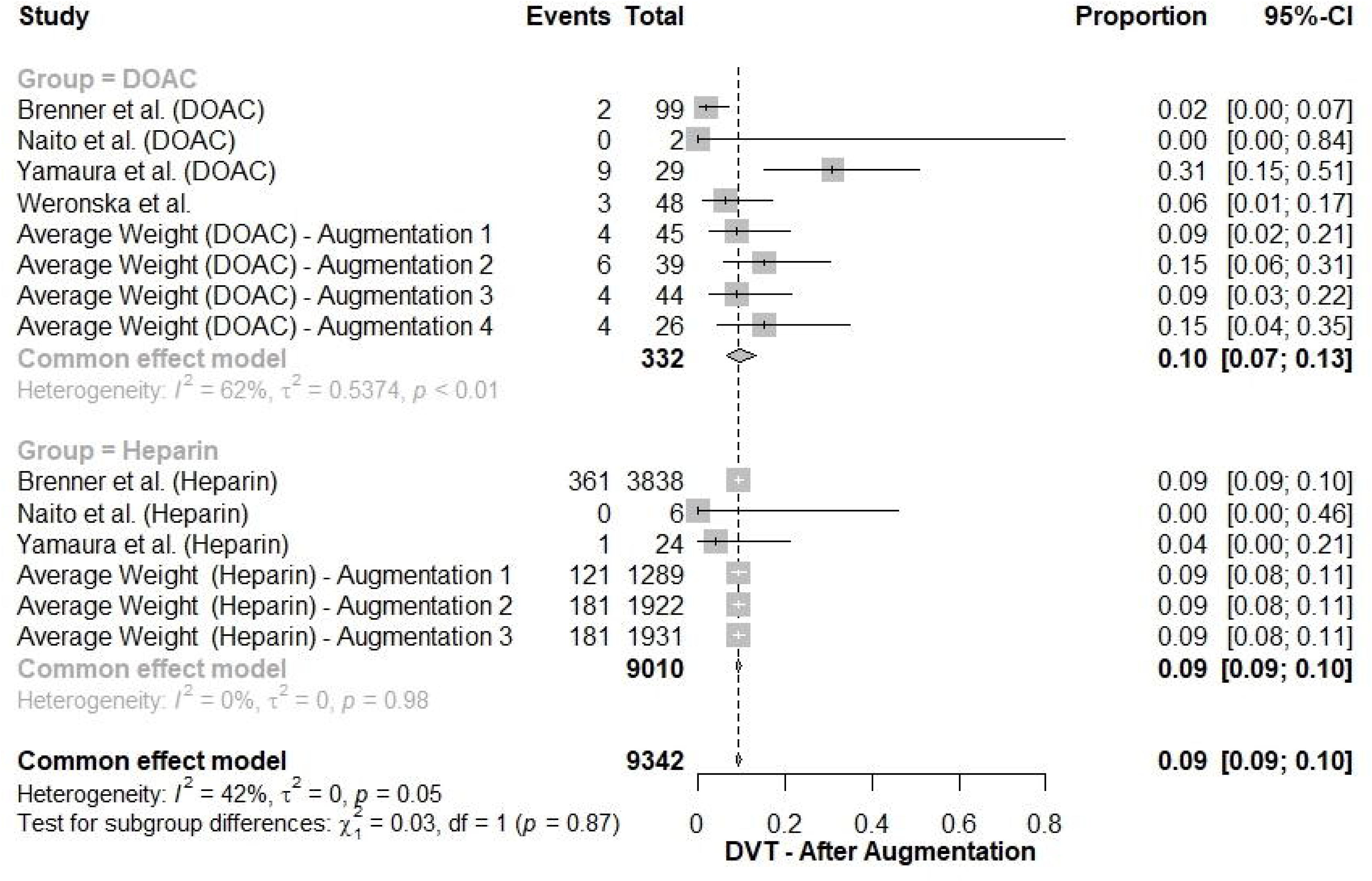
Sensitivity analysis to resolve heterogeneity in DVT rates for DOAC group after augmentation.

**Figure 6B:**
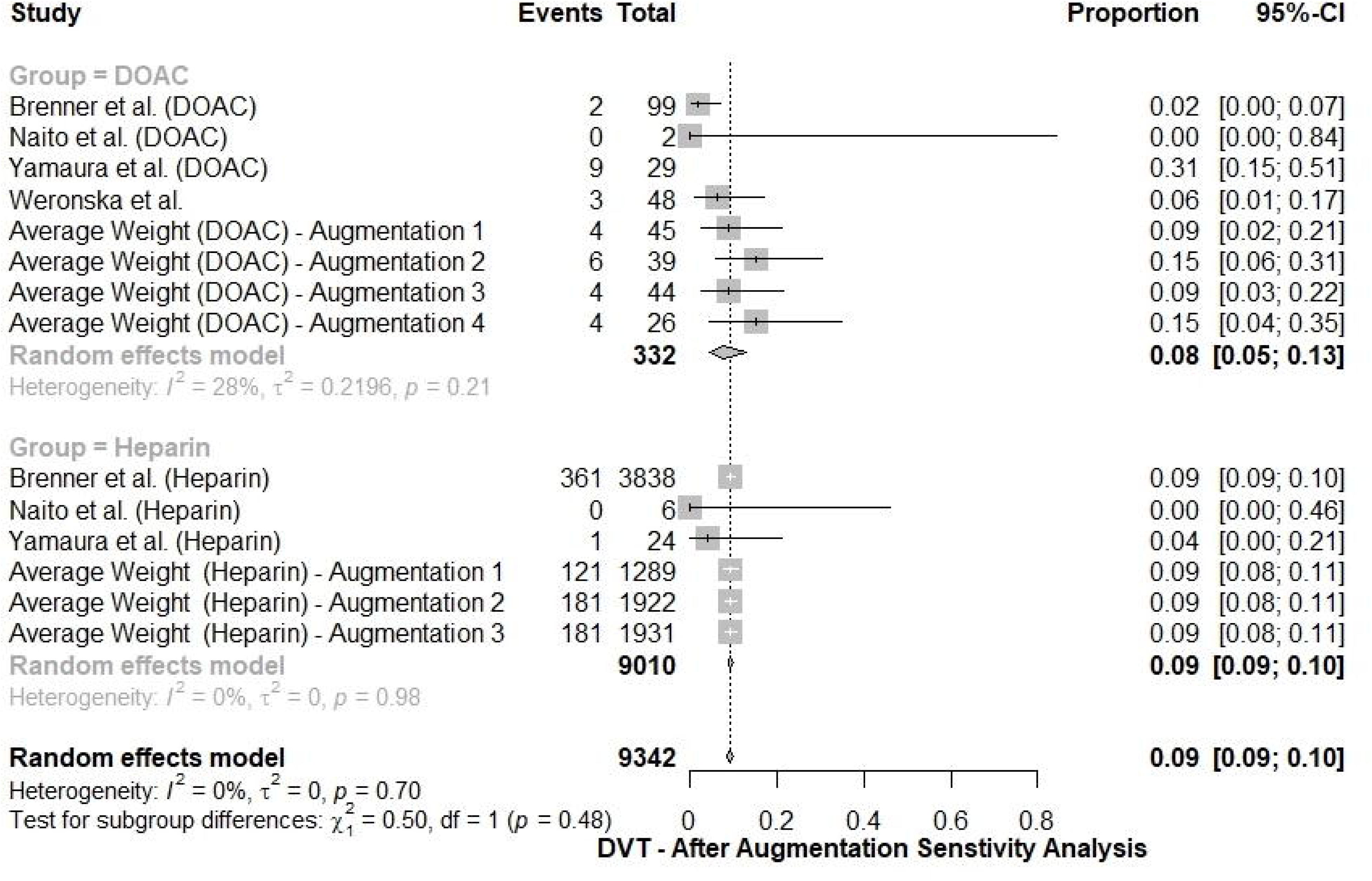
DVT rates for DOAC versus heparin after sensitivity analysis.

The clinically significant hemorrhage rate after augmentation was 4.0% (95% CI: 3-5%) with DOAC versus 9.0% (95% CI: 6-16%) with heparin. High heterogeneity existed for heparin (I2=79%, p<0.01). Sensitivity analysis did not resolve the heparin heterogeneity but did resolve the overall heterogeneity (I2=26%, p=0.15) (**Figure 7A**). The rate was lower with DOAC (4.0%, 95% CI: 3-5%) versus heparin (8.0%, 95% CI: 4-14%) (p=0.04) (**Figure 7B**).

**Figure 7A:**
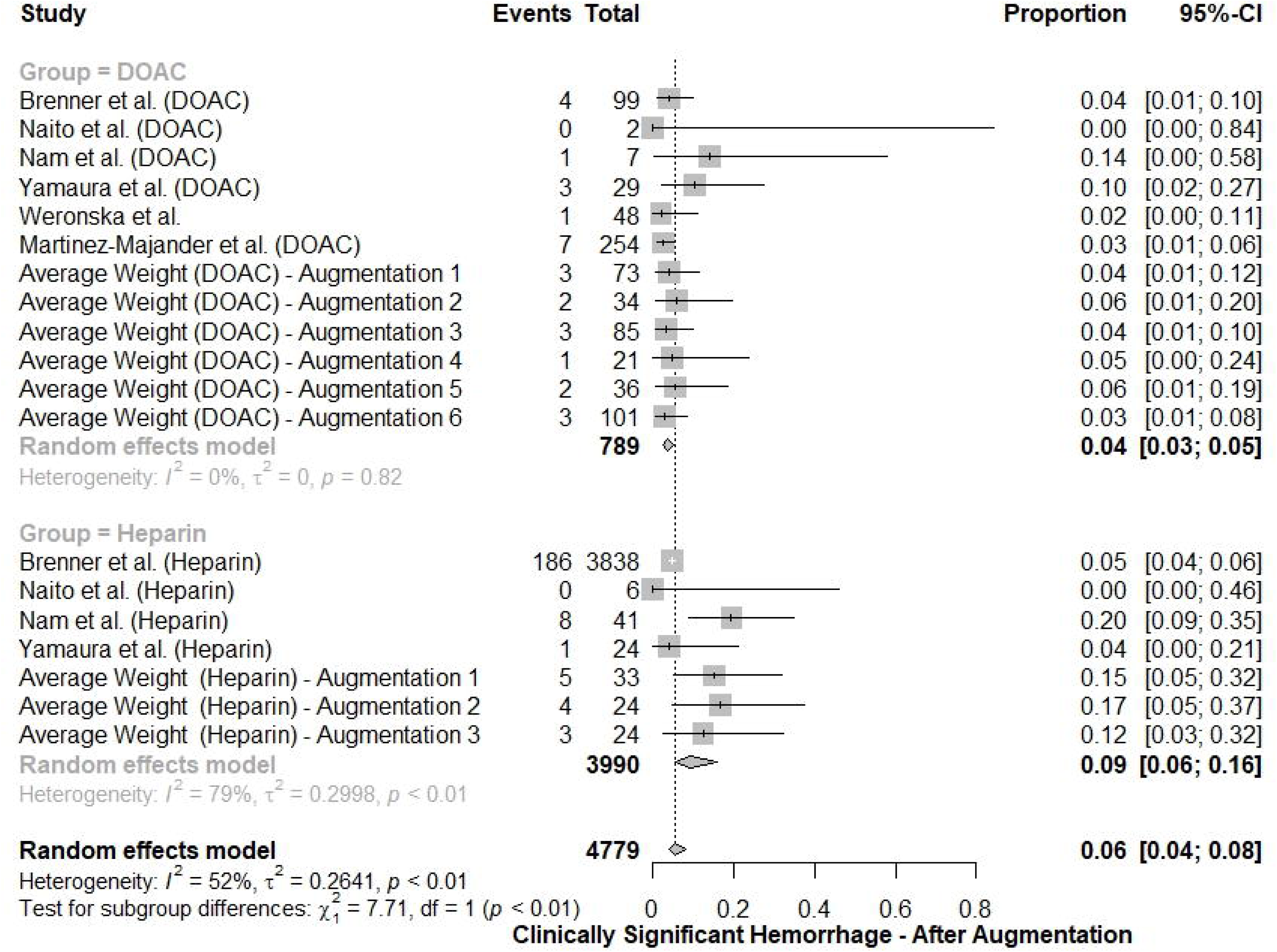
Sensitivity analysis to resolve heterogeneity in clinically significant hemorrhage rates, particularly for heparin group, after augmentation.

**Figure 7B:**
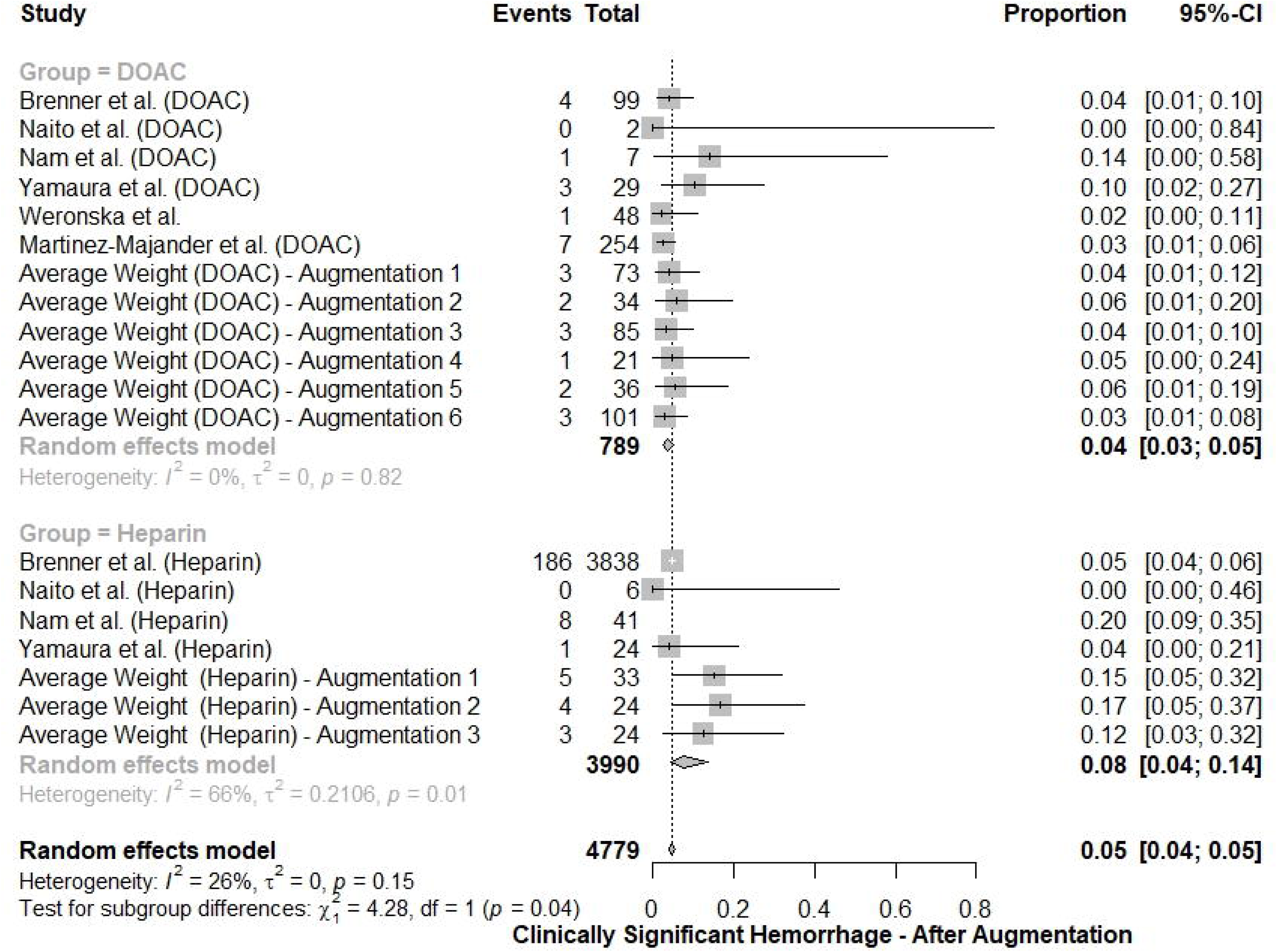
Clinically significant hemorrhage rates for DOAC versus heparin after sensitivity analysis.

The intracerebral hemorrhage rate after augmentation was 3.0% (95% CI: 1-7%) with DOAC versus 10.0% (95% CI: 6-17%) with heparin, with a significant difference (p=0.01) (**Figure 8**).

**Figure 8:**
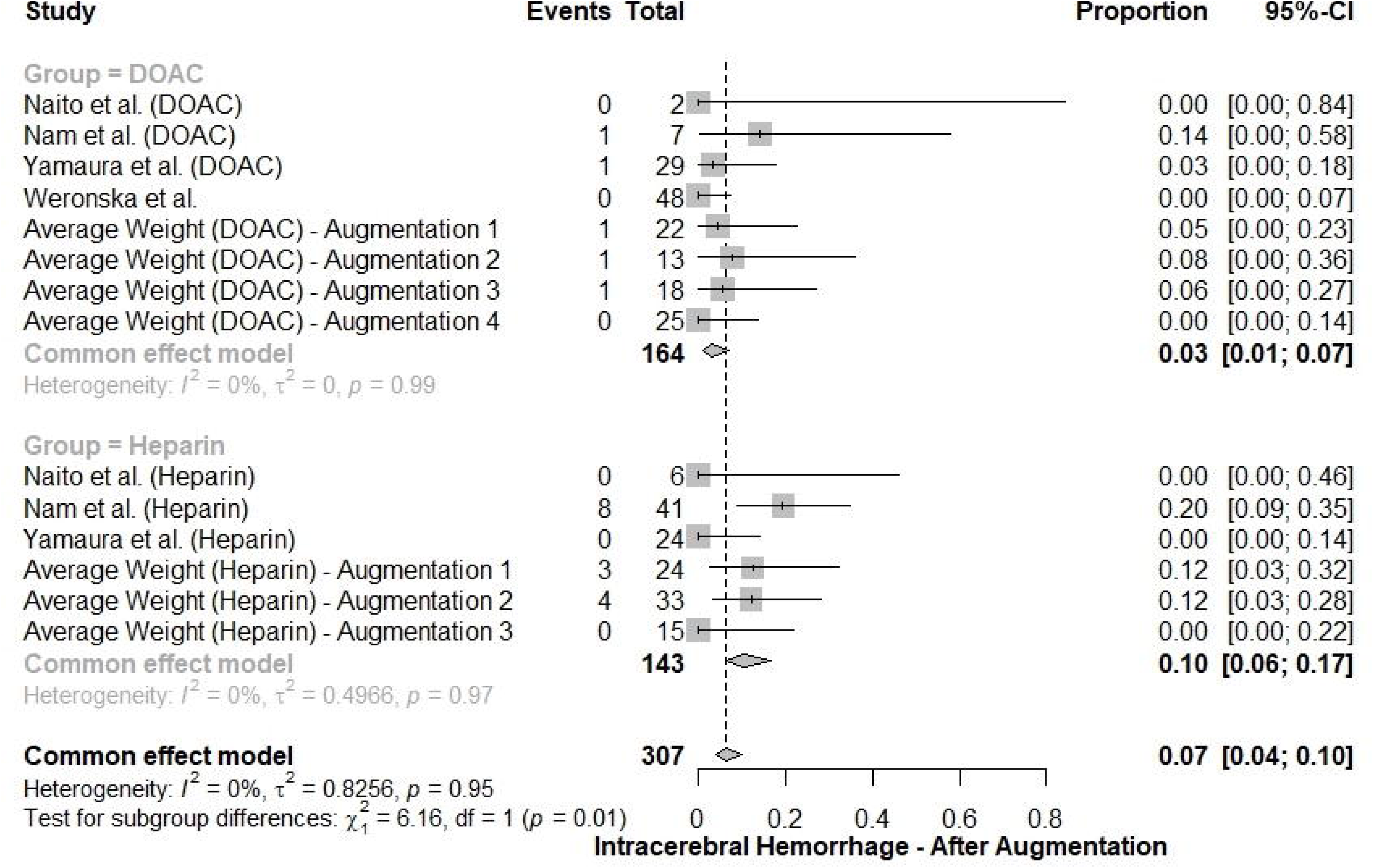
Intracerebral hemorrhage rates for DOAC versus heparin after augmentation.

The overall hemorrhagic complications rate after augmentation was 5.0% (95% CI: 3-7%) with DOAC versus 23.0% (95% CI: 16-31%) with heparin, with a significant difference (p<0.01) (**Figure 9**).

**Figure 9:**
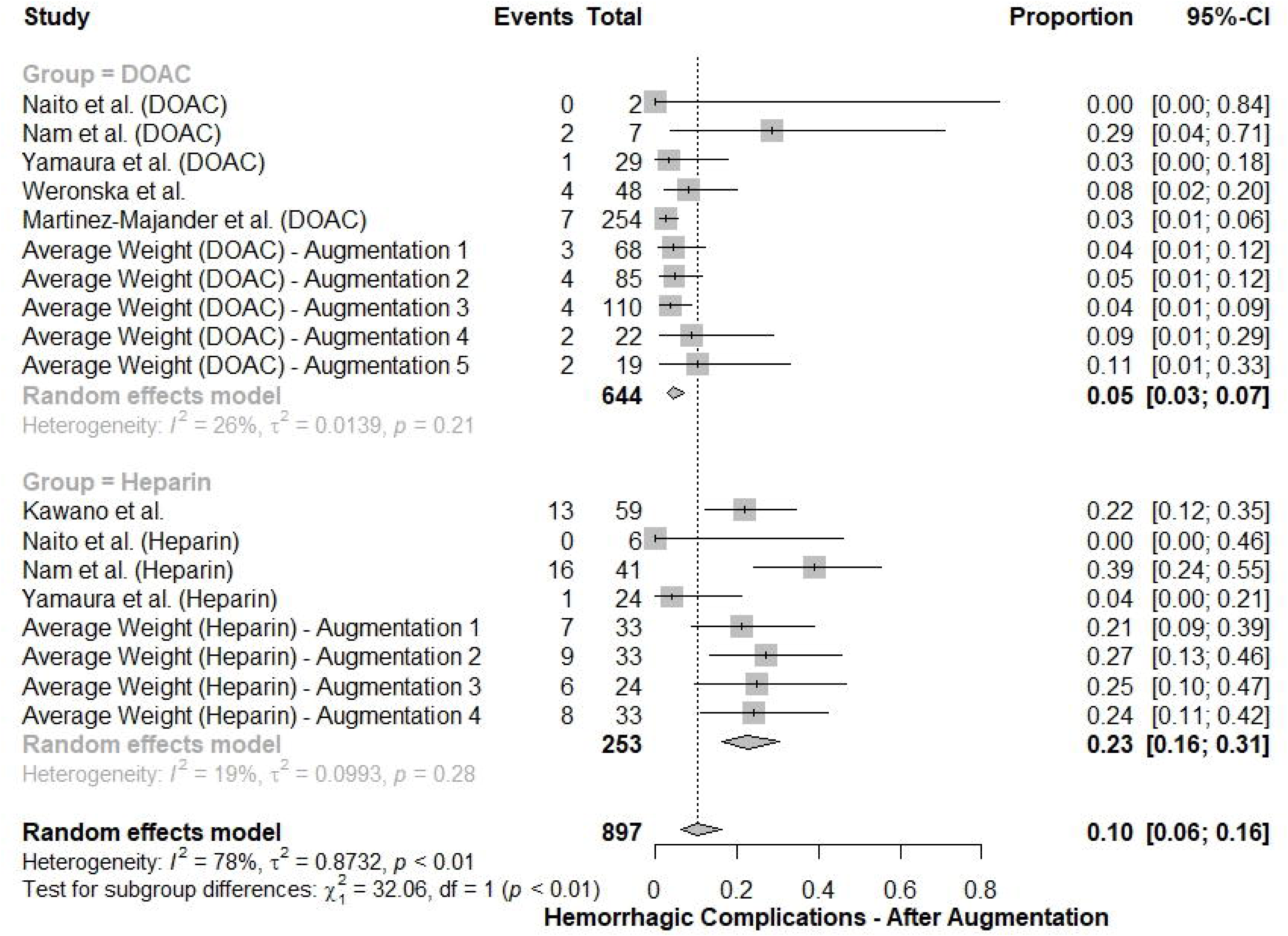
Overall hemorrhagic complications rates for DOAC versus heparin after augmentation.

The good clinical outcome rate (mRS 0-2) after augmentation was 39.0% (95% CI: 30-48%) with DOAC versus 27.0% (95% CI: 22-34%) with heparin, with a borderline difference (p=0.049) (**Figure 10**).

**Figure 10:**
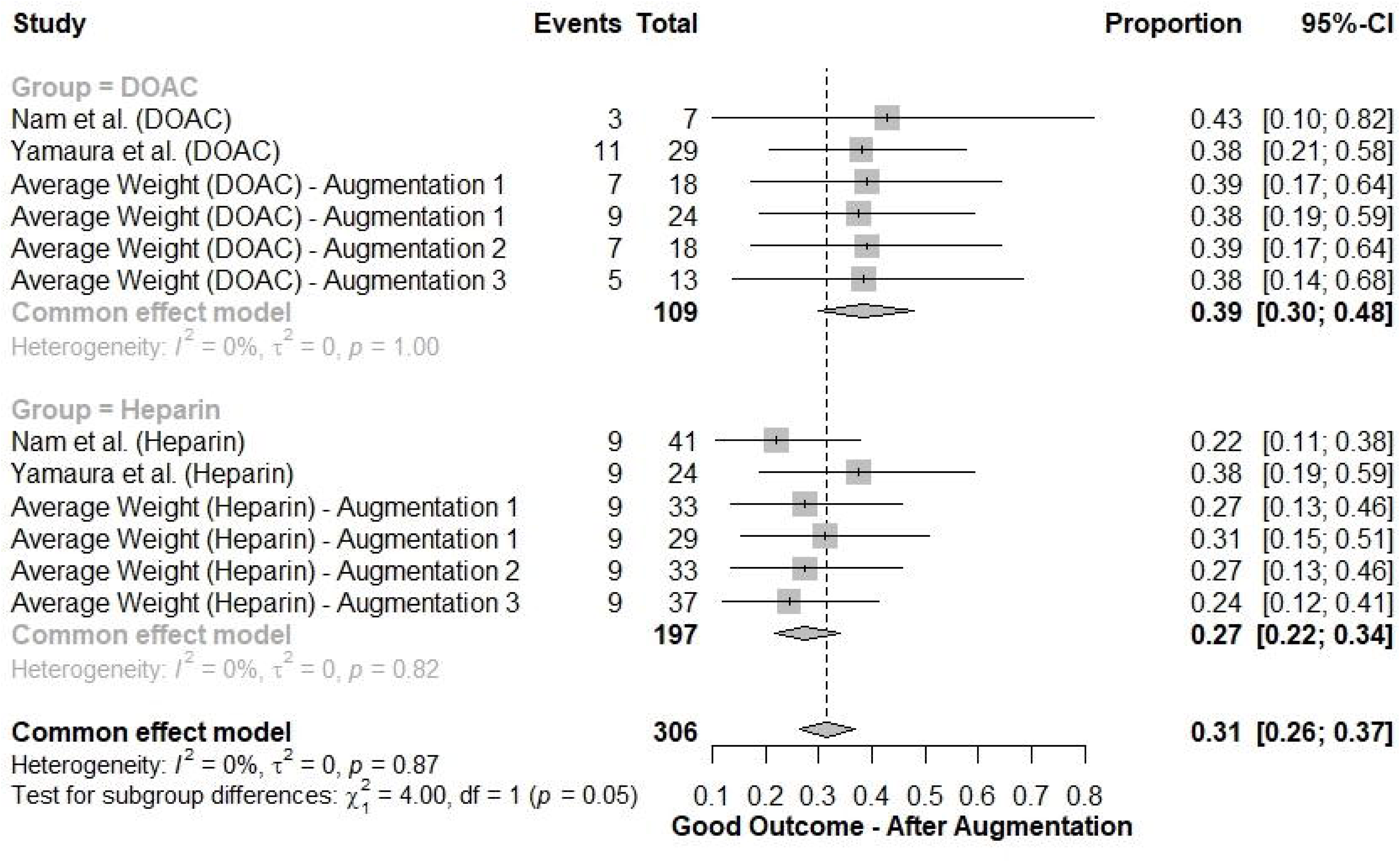
Good clinical outcome (mRS 0-2) rates for DOAC versus heparin after augmentation.

The mortality rate after augmentation was 12.0% (95% CI: 6-23%) with DOAC versus 33.0% (95% CI: 23-44%) with heparin. High heterogeneity existed for both groups. Sensitivity analysis resolved the heterogeneity (**Figure 11A**). The mortality rate was lower with DOAC (7.0%, 95% CI: 4-10%) versus heparin (27.0%, 95% CI: 20-35%), with a significant difference (p<0.01) (**Figure 11B**).

**Figure 11A:**
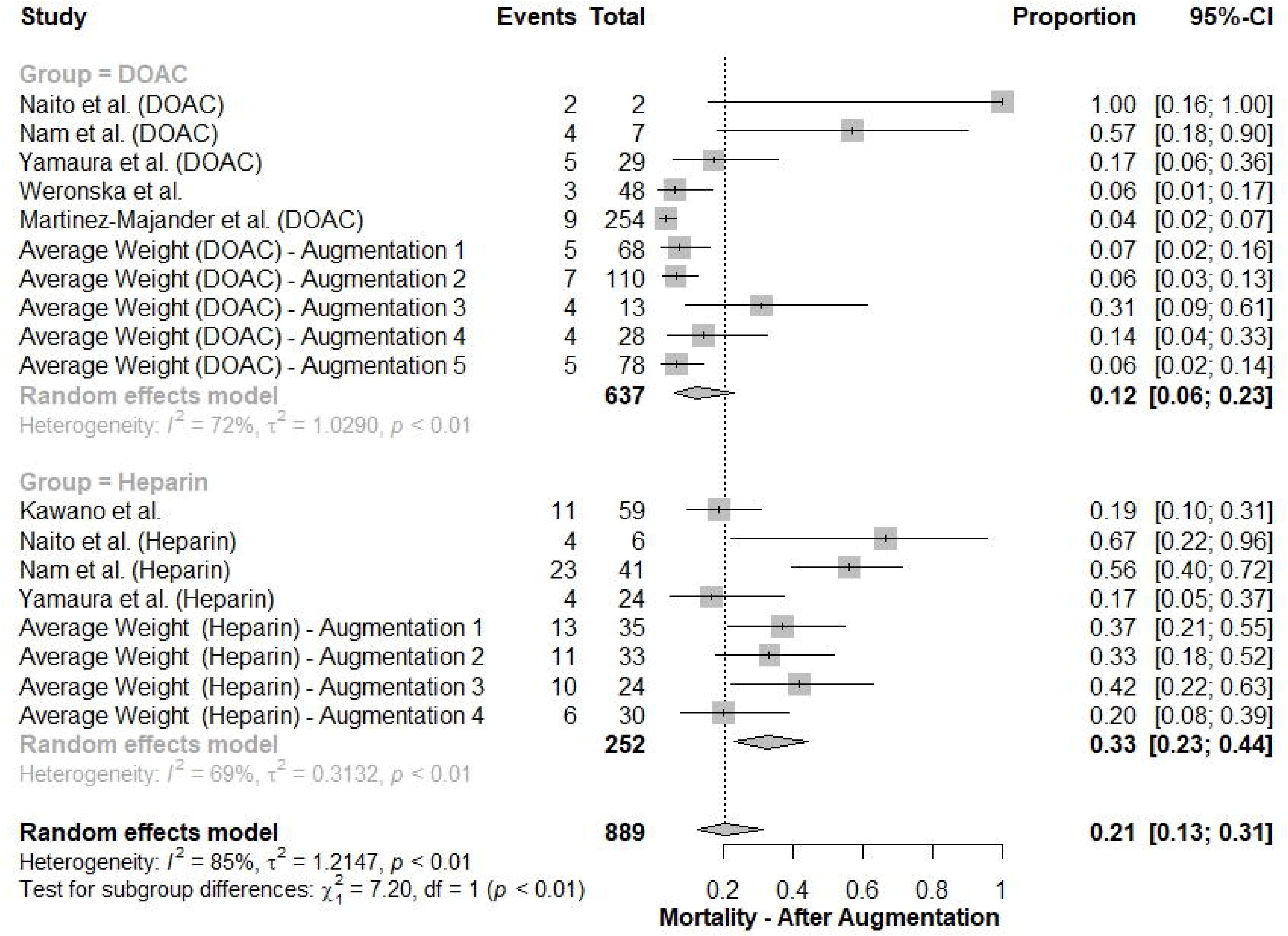
Sensitivity analysis to resolve heterogeneity in mortality rates for DOAC and heparin groups after augmentation.

**Figure 11B:**
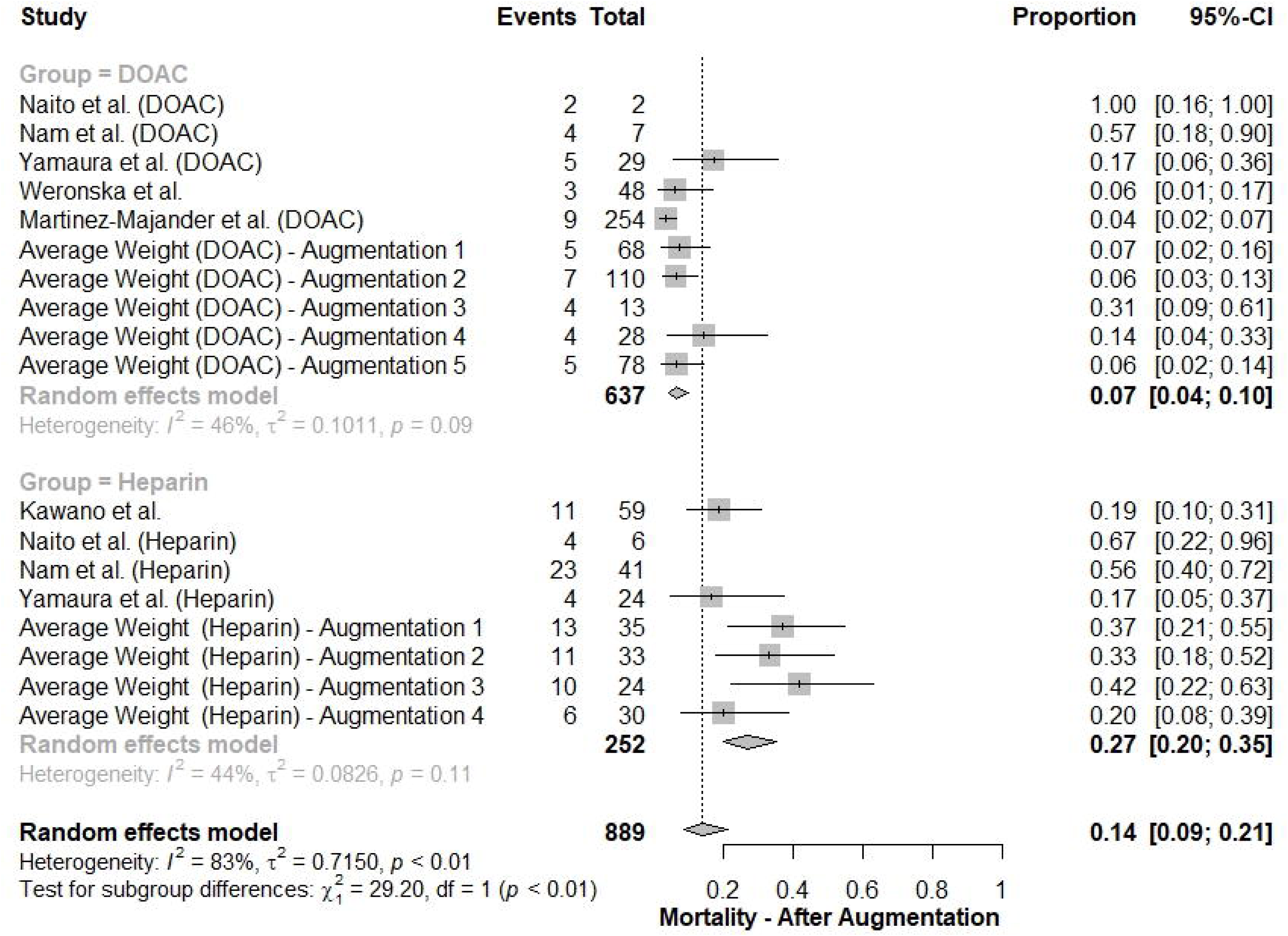
Mortality rates for DOAC versus heparin after conducting sensitivity analysis.

### Risk of bias

The risk of bias was low in two, moderate in two, and high in three studies (Supplemental Table 1). The largest study included 5717 patients, and the lowest included 26 patients. Among included studies, one was a randomized controlled trial (RCT)(26), one was a multiple-arm prospective multicenter cohort study(22), and the rest were retrospective observational studies; three were multiple-arm and two were single arm studies.

## DISCUSSION

Our meta-analysis of 4,407 patients provides a summary of the current data regarding safety and efficacy of DOACs and heparin-based products for the management of ischemic stroke related to systemic malignancy. Overall, 439 patients were treated with DOACs and 3,968 were treated with heparin-based products. There were significant differences in patient characteristics in terms of cancer type, most notably with hematologic malignancy more frequently reported among patients treated with DOACs. Intracerebral hemorrhage, and overall hemorrhagic complications were lower among patients treated with DOAC compared to heparin products. Rates of neurological outcomes, clinically significant hemorrhage, and recurrent ischemic stroke and venous thrombosis were similar among the two groups. All-cause mortality was more frequent in patients receiving heparin compared to DOACs. Machine learning based augmented meta-analysis showed similar results. These findings suggest DOACs have an acceptable safety and tolerability for secondary prevention of ischemic stroke in patients with malignancy, which is in line with previous reports showing a favorable safety profile of DOACs in other conditions (28, 29).

Despite frequent diagnosis of ischemic stroke in patients with systemic malignancy, optimal approach to secondary population remains unclear (2). This is in part due to the difficulty in establishing a stroke mechanism with many events described as ESUS (3). Use of DOACs compared to antiplatelet therapy in the setting of ESUS was the subject of two large clinical trials (13, 14). RESPECT-ESUS and NAVIGATE-ESUS showed no benefit from use of dabigatran or rivaroxaban compared to aspirin for the prevention of recurrent ischemic stroke in the setting of ESUS (13, 14). However, these studies did not specifically evaluate patients with underlying malignancy. The use of DOACs compared to heparin products in patients with ischemic stroke related to malignancy has not been prospectively evaluated. In the absence of high-quality prospective data from clinical trials, our meta-analysis supports the evidence on the safe use of DOACs in patients with ischemic stroke related to malignancy, demonstrating efficacy and safety profile compared to heparin products. Thus, this calls for longitudinal head-to-head cohort studies and future randomized controlled trials to further evaluate the use of DOACs for the treatment of ischemic stroke related to malignancy.

Although these observations stem from a limited number of patients with heterogenous cancer types in retrospective cohorts, they suggest that DOACs are potentially safer alternative to heparin, in terms of prevention of stroke and other thrombotic events in the setting of systemic malignancy. However, it is important to note that the smaller sample size of patients treated with DOACs, and the scarcity of incident events prevent us from arriving at a definitive conclusion. Therefore, it is prudent to interpret the study findings with caution and warrant further research with larger cohorts to validate these potential benefits.

### Limitations

The main limitation of this meta-analysis is the retrospective nature of most included studies with limited available data regarding baseline patient characteristics, type of malignancy diagnosed, and stroke mechanism. Heterogeneity in the design of studies included further added to the difficulty of result interpretation and aggregate analysis. Due to the small number of studies included, stratification by study design was not a possible solution. Incidences of solid organ tumors including gastric, colorectal, hepatobiliary, breast, and genitourinary cancer were also different among the patient groups. Furthermore, a higher proportion of metastatic disease and hematologic malignancy was observed in the DOAC group. This likely represents selection bias and limited data available regarding cancer types in the included retrospective studies. As such, stratification based on tumor type and proposed stroke mechanism was also not possible. Another limitation is the heterogeneity of indications for anticoagulation across studies and the lack of a clear definition of active cancer in two studies which leads to potential for interpretive errors. Relatively recent introduction of DOACs into clinical practice and paucity of prospective clinical trials studying ischemic stroke in patients with systemic malignancy explain the paucity of available data. Outcome measures including recurrent thrombotic events, hemorrhagic events, and neurological status were also not consistently available across the studies. The follow up periods were heterogeneous, making stroke outcome and all-cause mortality data difficult to interpret. Our study also does not address the question of how DOAC therapy may compare to antiplatelet therapy among patients with systemic malignancy experiencing an ischemic stroke. Finally, although newly-diagnosed cancer is accompanied by a significantly increased risk of incident AF and there is an overall higher incidence of AF in cancer patients,(30) we did not include patients with AF due to the inconsistency of the data, which adds further limitations to our study.

## CONCLUSION

DOACs are frequently prescribed for patients with hematologic malignancy or solid organ tumors who experience an ischemic stroke or TIA. In the absence of prospective clinical trial data, our meta-analysis of retrospective studies provides justification for the use of these agents in clinical practice with comparable safety and efficacy suggested. Given the small number of retrospective studies included and the limited data available, these findings should be interpreted with caution and require prospective validation.

## Data Availability

N/A

## Conflicts of Interest

N/A.

## IRB Approval

N/A.

## Funding Source

The project described was supported by the National Center for Advancing Translational Sciences (NCATS), National Institutes of Health, through CTSA award number: UM1TR004400. The content is solely the responsibility of the authors and does not necessarily represent the official views of the NIH.

## Ethical Approvals

N/A.

## Consent for Participation

N/A.

---

**Supplemental Table 1:** Assessment of risk bias in the included studies

