## Supplementary Table 1 for "Direct Oral Anticoagulants vs. Heparin for Cancer-Related Stroke: Augmented Meta-Analysis"

**Supplemental table 1:** Details of risk of bias assessment for included studies using ROBINS-I tool.

| Study (*et al*) | Year | Design | Confounding | Selection | Classification | Correlation with clinical practice | Missing data | Reported results |
| --- | --- | --- | --- | --- | --- | --- | --- | --- |
| Brenner et al. | 2018 | P, C, MC | Baseline characteristics were defined. Significant difference by baseline characteristics was reported. No control for confounding was performed. | Selection of patient for treatment was performed based on the treatment protocol of each hospital and at the discretion of the treating neurologist. Diagnosis criteria were well defined. Selection bias is low. | Treatment group was well defined. Prospective design. Probability of the classification of intervention by the outcome is very low. | Treatments available in the clinical practice were applied. Outcomes of the study are compatible with previous results of large RCTs. | No missing data were mentioned by the author. Data were available for all outcomes. | Outcome assessment was performed based on predefined guidelines. Potential influence by the previous knowledge of outcomes before publication is improbable due to the prospective design of the study. |
| Kawano et al | 2019 | R | Baseline characteristics were defined. No control for confounding was performed. | Selection of patient for treatment group was performed at the discretion of the treating neurologist. Diagnosis was performed based on DWI MRI. No information about the diagnosing radiologist/physician was provided. Selection bias is high. | Treatment group was well defined. Retrospective design. Probability of the classification of intervention by the outcome is very high. | Treatment available in the clinical practice was applied. | No missing data were mentioned by the author. Data were available for all outcomes. | Outcome assessment looks to be performed by the treating physician. Potential influence by the previous knowledge of outcomes before publication is possible due to the retrospective design of the study. |
| Martinez-Majander et al. | 2020 | RCT, C | A well performed RCT with major confounders controlled for. | Randomization with quadrable masking was performed. Selection and publication bias is improbable. Selection bias is improbable. | Treatment group was well defined. Prospective design. Probability of the classification of intervention by the outcome is very low. | Two treatment groups were available: Aspirin and DOAC. Both are available in clinical practice. Only DOAC group was included in this meta-analysis. | Control for missing data was performed. Most patients were available on last follow-up. | Potential influence by the previous knowledge of outcomes before publication is improbable due to the prospective study design, randomization of the cohort, and masking of the outcome assessor. |
| Naito et al. | 2018 | R, C | Baseline characteristics were defined. No control for confounding was performed. | No information about the treatment selection process was provided. Diagnosis of cancer related stroke was performed based on previous trial guidelines. No information about the stroke diagnosis neither the diagnosing radiologist/physician were provided. Selection bias is high. | Treatment group was well defined. Retrospective design. Probability of the classification of intervention by the outcome is very high. | Treatments available in the clinical practice were applied. | No missing data were mentioned by the author. Data were available for all outcomes. | Outcome assessment looks to be performed by the treating neurologist. Potential influence by the previous knowledge of outcomes before publication is possible due to the retrospective design of the study. |
| Nam et al. | 2017 | R, C | Baseline characteristics were defined. No significant difference in baseline characteristics between treatment was observed. No control for confounding was performed. | Selection of patient for treatment group was performed at the discretion of the treating neurologist. Diagnosis was performed based on DWI MRI by two independent neuroradiologists. Selection bias is uncertain. | Treatment group was well defined. Retrospective design. Probability of the classification of intervention by the outcome is very high. | Treatments available in the clinical practice were applied. | No missing data were mentioned by the author. Data were available for all outcomes. | Outcome assessment was performed by the treating neurologist and two independent neuroradiologists. Potential influence by the previous knowledge of outcomes before publication is possible due to the retrospective design of the study. |
| Weronska et al. | 2021 | R, C | Baseline characteristics were defined. No comparator treatment was available in this study based on the inclusion criteria of this meta-analysis No control for confounding was performed. | Selection of patient for treatment group was performed at the discretion of the treating neurologists. Diagnosis was performed using color duplex sonography. No information about the diagnosing radiologist/physician was provided. Selection bias is high. | Treatment group was well defined. Retrospective design. Probability of the classification of intervention by the outcome is very high. | Treatments available in the clinical practice were applied. | No missing data were mentioned by the author. Data were available for all outcomes. | Outcome assessment looks to be performed by the treating neurologist based on predefined guidelines. Potential influence by the previous knowledge of outcomes before publication is possible due to the retrospective design of the study. |
| Yamaura et al. | 2021 | R, C | Baseline characteristics were defined. No significant difference in baseline characteristics between treatment was observed. No control for confounding was performed. | Selection of patient for treatment group was performed at the discretion of the treating neurologist. Diagnosis was performed based on DWI MRI. No information about the diagnosing radiologist was provided. Selection bias is high. | Treatment group was well defined. Retrospective design. Probability of the classification of intervention by the outcome is very high. | Treatments available in the clinical practice were applied. | No missing data were mentioned by the author. Data were available for all outcomes. | Outcome assessment was performed by the two blinded neurologists and two independent neuroradiologists. Potential influence by the previous knowledge of outcomes before publication is possible due to the retrospective design of the study. |

**Abbreviations:** C, comparative (study); MC, multi-center study; P, prospective; R, retrospective, RCT, randomized clinical trial
